# Predicting Post-Traumatic Epilepsy with Automated Contusion Measurements using Acute CT Images: A Competing Risk Approach

**DOI:** 10.1101/2025.11.25.25341016

**Authors:** B. Berke Ayvaz, Justin R. Wheelock, Daniel S. Jin, Jenna Appleton, Samuel B. Snider, Victor Torres-Lopez, Neha Shrishail, William Sanders, Daniel Doherty, Khai Theeng Chow, Jisoo Kim, Maia Schlecter, Lawrence J. Hirsch, Adithya Sivaraju, M. Brandon Westover, Sahar F. Zafar, Aaron F. Struck, Kevin N. Sheth, Sacit Bulent Omay, Brian L. Edlow, Emily J. Gilmore, Jennifer A. Kim

## Abstract

**Objective:** This study evaluates contusion volume and location as predictors of PTE while considering mortality as a competing risk using automated segmentation of acute CT images.

**Methods:** Adult TBI patients who visited our center (2014-2025) were identified and categorized into three outcome groups: PTE, post-TBI mortality, and event-free. Clinical covariates were extracted from health records, and CT scans were processed using the BLAST-CT algorithm to segment intraparenchymal hemorrhage (IPH), edema, intraventricular (IVH), and extra-axial hemorrhage (EAH). Five-fold nested cross-validation was performed, with L1-regularized regression used for feature selection within each training fold and model performance evaluated in the corresponding test fold. Within each fold, the selected features were then used to fit separate cause-specific multivariable Cox models for PTE and mortality. Contusion location associations were assessed using both multivariable Cox and sparse canonical correlation analysis.

**Results:** Of 1017 identified patients, 6.1% developed PTE, 10.5% died, and 83.4% had no event. In multivariable analysis, total contusion volumes (p<0.001) were independently predicted both PTE and mortality, meanwhile and EAH volume (p<0.01) predicted mortality only. When regionalized, frontal (p<0.01) and temporal (p<0.01) contusions independently predicted PTE, whereas frontal (p=0.04) contusions also independently predicted mortality. Sparse canonical correlation analysis identified an association of frontotemporal and insular contusions with PTE but not mortality.

**Interpretation:** Automatically measured contusion segmentation from acute CT imaging could help predict post-traumatic epilepsy. A competing risk framework highlighted that contusion volume is associated with both PTE and mortality risk, whereas unique contusion location patterns may aid in post-traumatic epilepsy assessment in a broad TBI population.

## INTRODUCTION

Post-traumatic epilepsy (PTE), a long-term complication of traumatic brain injury (TBI), is characterized by unprovoked seizures occurring more than seven days after injury.^1,2^ PTE accounts for approximately 20% of structural epilepsies and 5% of all epilepsy cases.^3,4^ Despite decades of investigation, no anti-seizure medication has been shown to prevent PTE.^5,6^ Progress in therapeutic exploration has stalled due to high trial costs and the absence of reliable methods to identify individuals at high-risk.^7,8^ The unpredictable course of PTE in TBI survivors poses a major barrier to targeted prevention, but a reliable and generalizable prediction model could help overcome this by supporting early risk stratification and improving the feasibility of the trials.^9^

Previous efforts have identified biological, clinical, and neuroimaging-based biomarkers of PTE that reflect TBI burden.^7,10,11^ These findings have informed TBI classification^12^ and predictive models identifying patients at high risk of PTE.^13,14^ However, prior evidence often derives from specific research protocols or cohorts, limiting generalizability.^15–17^ Additionally, TBI carries mortality rates of up to 60% depending on severity, which may be of limited consideration in existing PTE models.^18,19^ Notably, post-TBI mortality and PTE share risk factors such as age, sex, and injury severity.^20–22^ Therefore, current predictive models may inadvertently introduce selection bias in their risk prediction assessment or not account for a competing outcome such as death.

The acute phase of TBI provides a key window to identify biomarkers and risk factors associated with PTE development. TBI presents with heterogeneous injury patterns,^12,23^ and their characterization with neuroimaging is essential. Cerebral contusions have been of particular interest in prognostic models, with larger contusions associated with worse outcomes.^14,24,25^ PTE studies have also examined contusion location in relation to increased risk.^13,16,26,27^ However, most contusion research currently relies on manual workflows, limiting large-scale implementation of such findings. Moreover, many studies utilize magnetic resonance imaging (MRI),^28,29^ which is not broadly available in acute TBI. Computed Tomography (CT) is the first line and most widely used imaging modality for acute TBI,^30^ yet most CT-based studies remain qualitative or require labor intensive manual segmentations.^13,26,27^ The recent development of BLAST-CT,^31^ an automated segmentation algorithm, enables rapid quantification of TBI-related lesions from standard clinical CT scans. This tool allows us to investigate associations between contusion volume and location with PTE and post-TBI mortality in scale.

The objective of this study was to use BLAST-CT to compute automated hemorrhage measurements in a large, clinically representative TBI cohort to: (1) predict PTE within a competing risk framework that accounts for post-TBI mortality, and (2) evaluate whether contusions in specific brain regions are associated with risk of PTE and post-TBI mortality.

## METHODS

In this study, data were collected from Yale Longitudinal Acute Brain Injury Biorepository and TBI Repository (New Haven, CT). The study protocol was approved by the Institutional Review Board (IRB #2000037022, #200031988) and granted a waiver of informed consent given the retrospective design.

### Study Cohort

Patients who had been treated at our tertiary care hospital between October 2014 and March 2025 were retrospectively identified. Adult (≥18 years) patients with TBI diagnosis and an available stability CT scan from the respective TBI admission were included. Individuals with any past medical history of seizures, epilepsy, brain malignancy, prior brain injury, or prior neurosurgical intervention that might confound our primary outcomes were excluded. Additional exclusions were applied for non-usable stability CT scans (e.g., missegmentation, corrupted images, or incomplete series) and missing features.

### Primary Outcomes

Primary outcomes included PTE, post-TBI mortality. Outcomes were identified via manual record review. PTE was defined as unprovoked clinical or electrographic seizure(s) occurring >7 days post-TBI.^32^ Post-TBI mortality was defined by identifying patients who were deceased at any time following the TBI event. Endpoints were defined as time from the TBI admission to the occurrence of the primary outcome, or to the latest known follow-up date for the patients without any primary event.

### Data Collection

For each patient, demographic information including age, sex, self-reported race, and ethnicity was collected. Clinical covariates were extracted by chart review, including Glasgow Coma Scale (GCS) with eye, verbal, and motor components; mechanism of traumatic brain injury; pupillary reactiveness; glucose and hemoglobin levels; any seizure event from the injury to the 7-day window; and any neurosurgical intervention, which included craniectomy, craniotomy, burr hole, intracranial monitoring device (e.g., intraparenchymal pressure, brain tissue oxygen monitor, depth electrode, etc.) or external ventricular drain within the first 7 days of the injury. For each patient, the first CT scan demonstrating hemorrhagic stability was identified and downloaded.

To collect quantitative imaging features, CT scans with axial slices of 5-mm thickness were used; if 5-mm slices were not available, 2.5-mm or 3-mm slices were used. Lesion segmentations were generated using the recently published automated segmentation algorithm, BLAST-CT.^31^ Though BLAST-CT was previously validated, its performance was further assessed for this study by comparing manually segmented contusion tracings from a preliminary cohort across two centers with automated BLAST-CT segmentations. BLAST-CT and manual contusion label volumes were correlated (R = 0.711; P < 0.0001) (Supplemental Fig. 1). For each scan, BLAST-CT automatically segments TBI lesions and creates a multi-class segmentation that includes intraparenchymal hemorrhage (IPH), intraparenchymal edema, intraventricular hemorrhage (IVH), and extra-axial hemorrhage (EAH). Nomenclature in both research and clinical practice recognizes trauma-caused intraparenchymal hemorrhages along a spectrum, and the terms (traumatic intracranial hemorrhage, contusion etc.) are often used interchangeably.^23^ We defined trauma-related IPH and edema complexes as contusions. Since contusions in CT scans can be composed of both hyperdense and hypodense areas, IPH and edema masks are combined to compute contusion volumes. For sensitivity analyses, a univariable analysis was performed in which IPH and edema volumes were evaluated separately (Supplemental Table 1).

An image processing pipeline was developed to compute atlas-based, regional contusion measurements using open-source tools.^33,34^ CT scans were first skull-stripped using ichseg,^35^ then registered to a 2-mm standard isotropic CT template using affine, rigid, and deformable registration.^36^ All registrations were manually inspected, and patients with misaligned atlas transformations were excluded. To allow the transformation of MNI atlas labels onto our patients’ CT spaces, the same registration was performed between the CT template and the MNI T1 1-mm template. Regional contusion volumes were calculated by overlapping BLAST-CT segmentations with all MNI atlas labels after transformation into patient CT space.^33,37^ All image processing was performed by BBA while blinded to outcome and adjudicated by JAK through visual inspection.

### Statistical Analysis

Demographic and clinical data were summarized using descriptive statistics with pairwise comparisons of outcome groups. Group differences in continuous variables were tested with ANOVA or Kruskal–Wallis based on Shapiro–Wilk normality test, and categorical variables were compared using χ² tests. Associations of individual features with outcomes were assessed with univariate Cox proportional hazards analyses volumes for post-TBI mortality and PTE. Univariate analyses were only used for descriptive purposes.

Two primary analytical models were developed: 1) a total hemorrhagic measurement model using total hemorrhagic segmentations and clinical features (Total Hemorrhage Model), and 2) a regional contusion model incorporating regionalized contusion measurements with the remaining total hemorrhagic measurements and clinical features (Regional Contusion Model). Additionally, a sensitivity analysis was performed after restricting the cohort to patients with a contusion present, either in isolation or in combination with other hemorrhagic lesions. Patients with contusions were identified by applying a ≥1 cc threshold to IPH and edema masks across the full cohort.

Separate cause-specific multivariable Cox proportional hazards models were constructed for PTE and post-TBI mortality, treating the competing event as censoring. Time-to-event was defined as the number of days from index TBI admission to the extracted time of the primary outcomes (PTE or post-TBI mortality), or to the last known follow-up date for the patients without any primary events. 5-fold nested cross-validation was performed for model evaluation and feature selection. Predictor selection was carried out using Least Absolute Shrinkage and Selection Operator (LASSO)–regularization within each training fold, and selected predictors were evaluated in the corresponding held-out test fold. Features selected in ≥60% of folds were retained for the final reported models. Model performance was derived from predictions on held-out test data and summarized as the average across five cross-validation folds. Model discrimination was quantified using the Harrell’s concordance index (C-index) and time-dependent area under the receiver operating characteristic curve (AUC), computed at fixed evaluation intervals from 6 to 36 months post-injury. Model calibration was assessed at each evaluation window by computing time-specific Brier scores to summarize prediction error over time. Calibration plots were also generated to compare predicted and observed event rates across quantile-based bins. Final models were retrained on the full dataset using the selected features from cross-validation. To visualize event probabilities over time, non-parametric cumulative incidence functions were estimated for PTE and mortality events using the Aalen–Johansen method, accounting for competing risks. To assess the incremental predictive contribution of imaging features, a model using only clinical features was fitted and compared with the total hemorrhage model using likelihood ratio test, detailed in Supplementary Materials.^38,39^

A voxelwise, multivariate lesion-to-symptom mapping analysis was then conducted to investigate how a given brain area is associated with each primary outcome where a contusion occurred. Contusion masks registered in CT template space, resampled to 3-mm voxel size and sparse canonical correlation analysis (SCCAN) was performed the LESYMAP package.^40,41^ SCCAN detects patterns of voxels that maximize the correlation between lesion patterns and the primary outcomes. Analysis included only voxels with lesions present in a minimum of 5 patients.^34,42^ The significance of the final weight map is assessed via a p-value computed from the Pearson correlation between voxel weights and outcomes. For each primary outcome, all patients were binarized into outcome versus non-outcome groups. Ultra-high-resolution MRI registered to MNI space was used for visualization of the resulting heatmap. Please see Snider et al. (2025) for further details on image processing and voxel-wise analysis.^34^

## RESULTS

### Cohort Characteristics

A total of 1,943 patients met the initial inclusion criteria. Exclusions due to ineligible medical history (n=418), unusable CT scans (n=285), and/or missing clinical features (n=363) resulted in a final cohort of 1,017 patients (Supplemental Figure 2). 6.7% of patients developed PTE, 10.5% died after TBI, and 83.4% had no event. The median time to PTE was 30 days (IQR:9–156), whereas the median time to death was 145 days (IQR:20–360). Baseline demographic and clinical characteristics of the cohort are detailed in Supplemental Table 2.

BLAST-CT segmentations were used to compute hemorrhage volumes. Median contusion volume was 0.50 cc (IQR 0.09–1.95), with a substantial proportion having near-zero measured hemorrhage. Median EAH volume was 0.37 cc (IQR 0.05–1.37). We performed a descriptive statistical analysis, stratified by outcome, and found that PTE patients had larger contusion volumes (6.12 cc, IQR:0.66–21.91) and EAH volumes (3.85 cc, IQR:0.58–12.03) compared with the other outcome groups. Post-TBI Mortality group had median contusion volumes of 1.07 cc (IQR 0.37–8.12) and EAH volumes of 0.97 cc (IQR 0.19–4.84) (Figure-1A). Additionally, patients were stratified by TBI severity based on GCS at admission. Median contusion volumes were larger in PTE patients across all severity subgroups. IVH volumes remained uniformly small across severity subgroups, though slightly higher in mortality patients across severity subgroups (Figure-1B, Supplemental Table 3).

**Figure 1.**
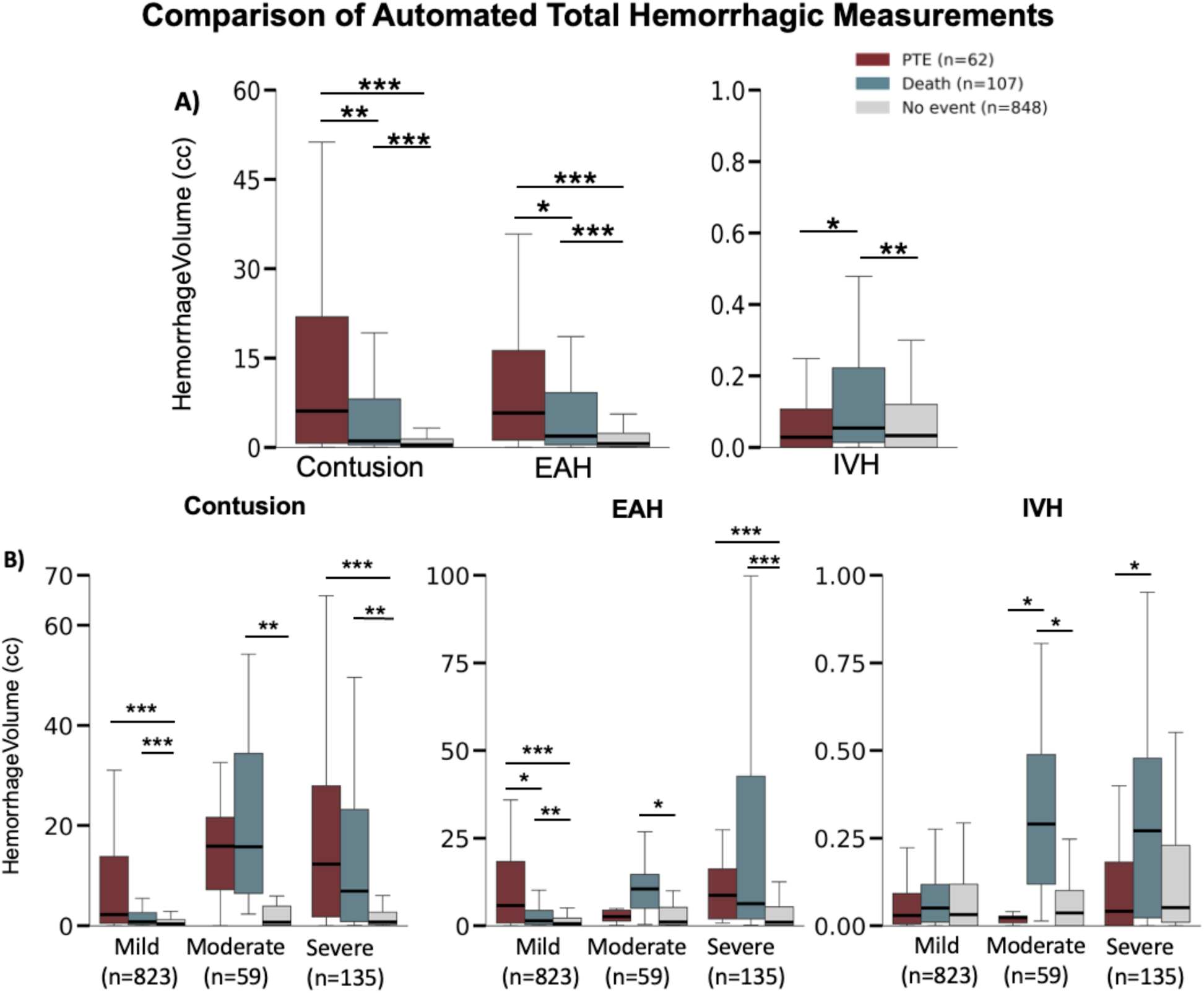
Distribution of automated hemorrhage volume measurements across PTE (red), mortality (blue), no event (gray) outcomes. A) Distribution of total lesion volumes for contusions (combined intraparenchymal hemorrhage and edema), extra-axial hemorrhage (EAH), and intraventricular hemorrhage (IVH) stratified by clinical outcome group (PTE, death, no event). B) Stratified distributions of contusion, extra-axial hemorrhage (EAH), and intraventricular hemorrhage (IVH) volumes across TBI severity groups (mild, moderate, severe) based on admission Glasgow Coma Scale scores. Statistical comparisons performed using Mann-Whitney U tests for pairwise comparisons between outcome groups within each severity stratum. Complete statistical comparison provided in Supplementary Table 3. *p<0.05, **p<0.01, ***p<0.001

Regionalized contusion volumes were computed and analyzed descriptively (Supplemental Table 4). Frontal and temporal areas showed slightly larger median contusion volumes, likely reflecting the predilection of contusions to involve these regions. Frontal contusions measured 1.40 cc (IQR 0.19–7.14) in PTE patients vs. 0.35 cc (IQR 0.04–2.30) in those who died. Temporal contusions followed a similar pattern, with medians of 0.71 cc (IQR 0.11–4.98) and 0.26 cc (IQR 0.02–1.10), respectively (Figure 2).

**Figure 2.**
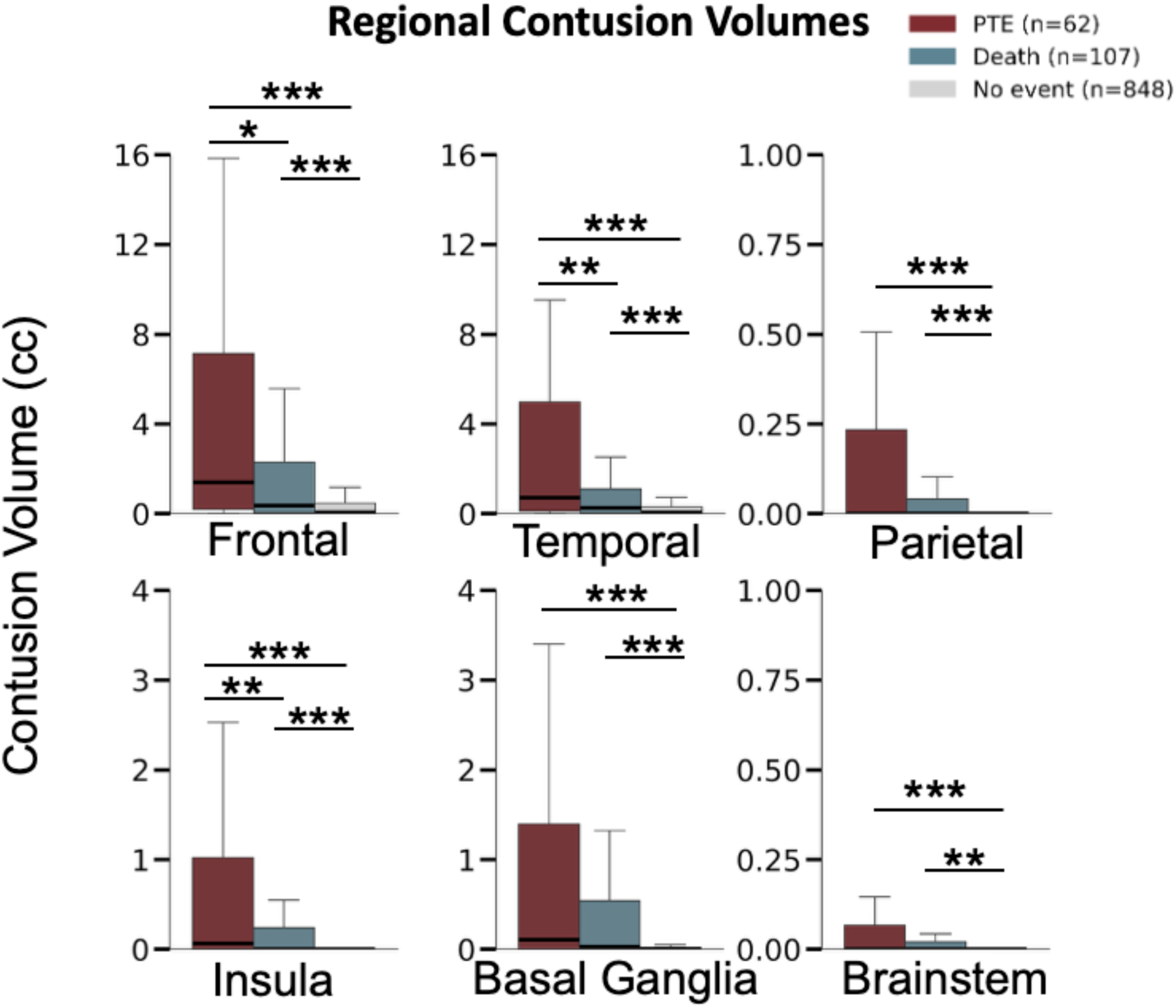
Regional contusion volume distributions across outcome groups. Boxplots showing regional lesion volumes (frontal, temporal, parietal, occipital, insular, basal ganglia, brainstem, and cerebellar contusions) stratified by outcome group (PTE, death, no event). Statistical comparisons performed using Mann-Whitney U tests for pairwise comparisons between outcome groups. Complete statistical comparison provided in Supplementary Table 4. *p<0.05, **p<0.01, ***p<0.001

### Univariable Analysis

Univariate cause-specific Cox proportional hazards models identified demographic, clinical, and neuroimaging variables significantly associated with PTE and post-TBI mortality (Table-1). Male sex was associated with greater PTE risk (HR 2.12, 95% CI 1.17–3.84) but not post-TBI mortality. Younger age was associated with PTE (HR 0.97, 95% CI 0.96–0.99) while older age associated with post-TBI mortality (HR 1.05, 95% CI 1.04–1.06). Greater injury severity was associated with both PTE and mortality. Laboratory values at admission also showed outcome-specific associations, such as a small but statistically significant association of increased blood glucose with PTE (HR 1.003, 95% CI 1.001–1.01), and lower hemoglobin with mortality (HR 0.73, 95% CI 0.67–0.79). Post-traumatic seizures (HR 7.54, 95% CI 4.45–12.76) and neurosurgical intervention within 7 days (HR 4.61, 95% CI 2.76–7.73) were strongly associated with increased PTE risk.

BLAST-CT derived measurements of contusion and EAH volumes were associated with increased risk of both PTE and mortality. Contusion volume was associated with increased risk of both PTE (HR_Δ5cc_ 1.20, 95% CI 1.15–1.25) and mortality (HR_Δ5cc_ 1.82, 95% CI 1.45–2.29). Similarly, EAH volume predicted PTE (HR_Δ5cc_ 1.15, 95% CI 1.10–1.21) and mortality (HR_Δ5cc_ 1.93, 95% CI 1.61–2.32). IVH volume was associated with mortality only (HR_Δ1cc_ 1.48, 95% CI 1.13-1.93). When regionalized, contusions at frontal, temporal, parietal, insular, basal ganglia and brainstem showed association with both outcomes, meanwhile occipital contusions were not associated with either outcome.

### Multivariable Analysis

#### Model 1: Total Hemorrhage Model

The multivariable Total Hemorrhage Model was developed for both PTE and post-TBI mortality by combining retained total hemorrhage volumes with demographic and clinical features. (Table 2). Several demographic and clinical features were independently associated with outcomes. Early post-traumatic seizures (HR 3.5, 95% CI 1.97-6.17), neurosurgical intervention within 7 days (HR 2.45, 95% CI 1.39-4.33), and lower GCS (HR 0.91, 95% CI 0.86-0.96) were independent predictors of post-traumatic epilepsy (PTE). Older age (HR 1.05, 95% CI 1.04-1.07), lower GCS (HR 0.86, 95% CI .81-0.90) and lower hemoglobin (HR 0.75, 95% CI 0.68-0.83) were independent predictors of mortality.

**Table 1.** Univariate cause-specific Cox proportional hazards models for PTE and post-TBI death.

| Features | PTE |  | Death |  |
| --- | --- | --- | --- | --- |
|  | HR (95%CI) | p | HR (95%CI) | p |
| **Demographics** |  |  |  |  |
| Age | 0.97 (0.96-0.99) | **<0.001** | 1.05 (1.04-1.06) | **<0.001** |
| Sex (Male) | 2.12 (1.17-3.84) | **0.013** | 0.78 (0.53-1.14) | 0.2 |
| **Clinical Features** |  |  |  |  |
| Total GCS | 0.85 (0.81-0.89) | **<0.001** | 0.94 (0.9-0.98) | **0.01** |
| GCS Eye Score | 0.57 (0.48-0.68) | **<0.001** | 0.83 (0.71-0.98) | **0.03** |
| GCS Verbal Score | 0.64 (0.56-0.74) | **<0.001** | 0.87 (0.77-0.98) | **0.02** |
| GCS Motor Score | 0.69 (0.61-0.77) | **<0.001** | 0.83 (0.71-0.98) | **0.03** |
| Pupillary Reaction (ref. Both reactive) |  |  |  |  |
| One | 0.7 (0.17-2.88) | 0.627 | 1.2 (0.53-2.73) | 0.67 |
| None | 2.77 (1.19-6.42) | 0.627 | 2.54 (1.28-5.03) | **0.01** |
| **Injury Mechanism (ref. GLF)** |  |  |  |  |
| Accel/Decel | 4.68 (2.81-7.8) | **<0.001** | 0.67 (0.34-1.33) | 0.25 |
| Direct Impact | 0.47 (0.23-0.95) | **0.035** | 0.46 (0.27-0.79) | **<0.001** |
| Fall > 3ft | 0.64 (0.26-1.59) | 0.334 | 0.77 (0.4-1.48) | 0.43 |
| Penetrating Injury<sup>a</sup> | NA | 0.995 | NA | 0.99 |
| Blood Glucose at Admission | 1.003 (1.001-1.006) | **0.001** | 1.0 (0.99-1.002) | 0.77 |
| Hemoglobin at Admission | 0.92 (0.81-1.04) | 0.181 | 0.73 (0.67-0.79) | **<0.001** |
| Posttraumatic Seizures <7 days | 7.54 (4.45-12.76) | **<0.001** | 1.05 (0.49-2.25) | 0.91 |
| Neurosurgical Intervention <7 days | 4.61 (2.76-7.73) | **<0.001** | 0.77 (0.4-1.47) | 0.43 |
| **Neuroimaging Variables** |  |  |  |  |
| **Total Segmentation Volumes** |  |  |  |  |
| IPH<sub>Δ5cc</sub> | 1.41 (1.31 – 1.52) | **<0.001** | 2.56 (1.51 – 4.35) | **<0.001** |
| IVH<sub>Δ1cc</sub> | 0.96 (0.42-2.18) | 0.922 | 1.48 (1.13-1.93) | **<0.001** |
| EAH<sub>Δ5cc</sub> | 1.16 (1.10 – 1.21) | **<0.001** | 1.94 (1.61 – 2.33) | **<0.001** |
| Edema<sub>Δ5cc</sub> | 1.34 (1.25 – 1.43) | **<0.001** | 2.97 (2.06 – 4.29) | **<0.001** |
| Contusion (IPH+Edema)<sub>Δ5cc</sub> | 1.20 (1.16 – 1.25) | **<0.001** | 1.82 (1.45 – 2.30) | **<0.001** |
| **Regional Contusion Volumes** |  |  |  |  |
| Frontal<sub>Δ1cc</sub> | 1.05 (1.04–1.07) | **<0.001** | 1.04 (1.02–1.06) | **<0.001** |
| Temporal<sub>Δ1cc</sub> | 1.10 (1.07–1.12) | **<0.001** | 1.05 (1.02–1.08) | **0.002** |
| Parietal<sub>Δ1cc</sub> | 1.08 (1.03–1.13) | **0.001** | 1.07 (1.02–1.12) | **0.005** |
| Occipital<sub>Δ1cc</sub> | 1.04 (0.92–1.19) | 0.526 | 1.04 (0.96–1.14) | 0.341 |
| Insular<sub>Δ1cc</sub> | 1.58 (1.38–1.81) | **<0.001** | 1.36 (1.17–1.57) | **<0.001** |
| Basal Ganglia<sub>Δ1cc</sub> | 1.10 (1.07–1.14) | **<0.001** | 1.06 (1.02–1.10) | **0.004** |
| Brainstem<sub>Δ1cc</sub> | 1.32 (1.09–1.60) | **0.004** | 1.38 (1.17–1.62) | **<0.001** |

PTE = post-traumatic epilepsy; HR = hazard ratio; CI = confidence interval; GCS = Glasgow Coma Scale; GLF = ground-level fall; IPH = intraparenchymal hemorrhage; IVH = intraventricular hemorrhage; EAH = extra-axial hemorrhage.

**Table 2.** Multivariable cause-specific Cox regression of the Total Hemorrhage Model (Model 1) for PTE and post-TBI mortality.

| Features | Coefficient (95%CI) | p-value |
| --- | --- | --- |
| **PTE** |  |  |
| Total Contusion Volume$\Delta_{5cc}$ | 1.17 (1.12-1.23) | **<0.001** |
| Post-traumatic Seizures <7 days | 3.5 (1.97-6.17) | **<0.001** |
| Neurosurgical Intervention <7 days | 2.45 (1.39-4.33) | **0.002** |
| Total GCS | 0.91 (0.86-0.96) | **0.001** |
| **Death** |  |  |
| Age | 1.05 (1.04-1.07) | **<0.001** |
| EAH Volume$\Delta_{5cc}$ | 1.08 (1.04-1.13) | **<0.001** |
| Total Contusion Volume$\Delta_{5cc}$ | 1.11 (1.05-1.17) | **<0.001** |
| Total GCS | 0.86 (0.81-0.90) | **<0.001** |
| Hemoglobin | 0.75 (0.68-0.83) | **<0.001** |

PTE = post-traumatic epilepsy; CI = confidence interval; GCS = Glasgow Coma Scale; GLF = ground-level fall; EAH = extra-axial hemorrhage; IVH = intraventricular hemorrhage.

Automated hemorrhagic measurements improved predictive performance for both outcomes. In the PTE model, total contusion volume (HR_Δ5cc_ 1.17, 95% CI 1.12-1.23) was independent predictor. In the mortality model, total contusion volume (HR_Δ5cc_ 1.11, 95% CI 1.05-1.17) and EAH volume (HRΔ5cc 1.08, 95% CI 1.04–1.13) were independently associated with increased risk. Using 5-fold cross-validation, the average concordance index (C-index) for the PTE model was 0.799, the mean AUC was 0.787 and mean Brier score was 0.056 across folds. The mortality model demonstrated an average C-index of 0.785, mean AUC of 0.808, and mean Brier score of 0.084 (Figure 3A; Supplemental Tables 5–6; Supplemental Figure 3A).

**Figure 3.**
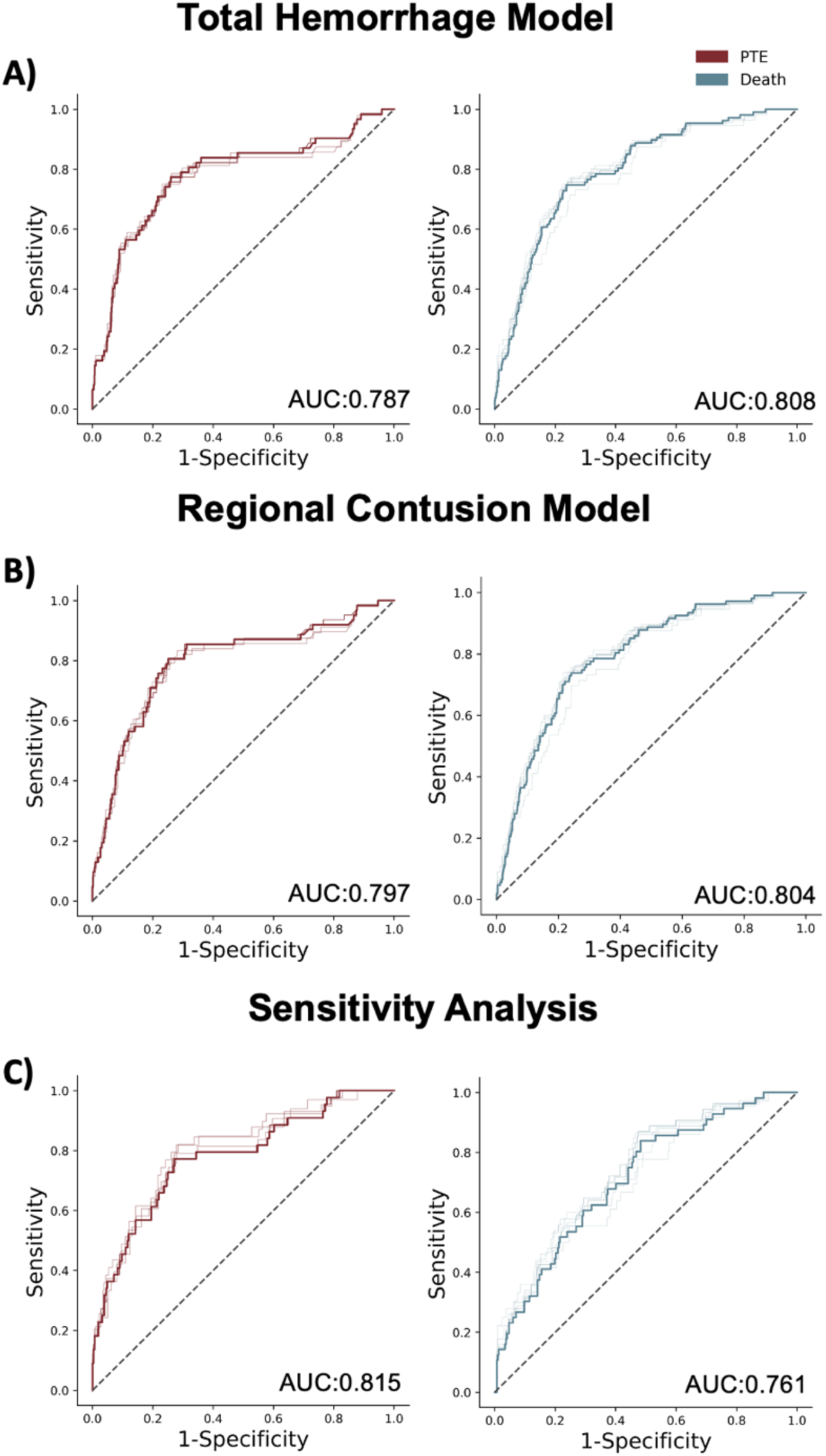
Receiver operating characteristic (ROC) curves for model discrimination. (A) Total Hemorrhage Model (Model 1) applied to the full TBI cohort (PTE mean time-dependent AUC = 0.787; Mortality = 0.808). (B) Regional Contusion Model (Model 2) incorporating regionalized contusion volumes with total EAH and IVH volumes (PTE mean time-dependent AUC = 0.797; Mortality = 0.804). (C) Sensitivity analysis restricted to patients with detectable contusions on BLAST-CT (PTE mean time-dependent AUC = 0.815; Mortality = 0.761). ROC curves illustrate discrimination for prediction of post-traumatic epilepsy (PTE) and post-TBI mortality over 6–36 months. Detailed timepoint-specific AUC values, calibration and performance metrics are provided in Supplemental Materials.

#### Model 2: Regional Contusion Model

To investigate the association of contusions within specific brain regions with outcomes, multivariable cause-specific Cox models were developed incorporating regionalized contusion volumes together with EAH, IVH, demographic and clinical covariates (Table 3). Independent predictors of PTE included neurosurgical intervention (HR 2.3, 95% CI 1.31-4.02), early post-traumatic seizures within 7 days (HR 3.41, 95% CI 1.89-6.18) and lower GCS (HR 0.91, 95% CI 0.86-0.96). Independent clinical predictors of mortality included older age (HR 1.06, 95% CI 1.04-1.07), lower GCS (HR 0.87, 95% CI 0.82-0.92), lower hemoglobin (HR 0.76, 95% CI 0.69-0.85) and no pupillary reaction at admission (HR 2.72, 95% CI 1.21-6.08).

**Table 3.** Multivariable cause-specific Cox regression of the Regional Contusion Model (Model 2) for PTE and post-TBI mortality.

| Features | Coefficient (95%CI) | p-value |
| --- | --- | --- |
| **PTE** |  |  |
| Post-traumatic Seizures <7 days | 3.41 (1.89-6.18) | **<0.001** |
| Neurosurgical Intervention <7 days | 2.3 (1.31-4.02) | **0.003** |
| Temporal Contusion Volume$\Delta_{1cc}$ | 1.05 (1.03-1.08) | **<0.001** |
| Frontal Contusion Volume$\Delta_{1cc}$ | 1.04 (1.02-1.07) | **<0.001** |
| Total GCS | 0.91 (0.86-0.96) | **0.001** |
| **Death** |  |  |
| Pupillary Reaction: None | 2.72 (1.21-6.08) | **0.01** |
| EAH Volume$\Delta_{5cc}$ | 1.08 (1.03-1.12) | **<0.001** |
| Frontal Contusion Volume$\Delta_{1cc}$ | 1.03 (1.01-1.06) | **<0.001** |
| Age | 1.06 (1.04-1.07) | **<0.001** |
| Total GCS | 0.87 (0.82-0.92) | **<0.001** |
| Hemoglobin at Admission | 0.76 (0.69-0.85) | **<0.001** |

PTE = post-traumatic epilepsy; CI = confidence interval; GCS = Glasgow Coma Scale; GLF = ground-level fall; EAH = extra-axial hemorrhage; IVH = intraventricular hemorrhage.

Automated measurement of frontal (HR_Δ1cc_ 1.04, 95% CI 1.02-1.07) and temporal (HR_Δ1cc_ 1.05, 95% CI 1.03-1.08) contusions were significantly associated with PTE. In the mortality prediction model, frontal contusion volume (HR_Δ1cc_ 1.03, 95% CI 1.001-1.06) and EAH volume (HR_Δ5cc_ 1.08, 95% CI 1.03–1.12) were significantly associated. Model performance was similar to the total hemorrhage model. For PTE, the regional model achieved an average C-index of 0.809, with a mean AUC of 0.797 and a mean Brier score of 0.057 across folds. For mortality, the model achieved an average C-index of 0.780, with an overall mean AUC of 0.804 and a mean Brier score of 0.084 (Figure 3B; Supplemental Tables 7–8; Supplemental Figure 3B).

#### Sensitivity Analysis

To analyze how restricting analyses to only patients who have at least one contusion (“contusion-inclusive”) might yield different findings compared with our broader cohort which includes TBI patients without contusions, a sensitivity analysis was performed. A cohort of 353 patients with contusions was identified, of whom 12.5% developed PTE, vs 6.7% in the broader cohort; 15.9% died after TBI, vs 10.5% in the broader cohort; and 71.7% had no event, vs 83.4% in the broader cohort (Figure 4).

**Figure 4.**
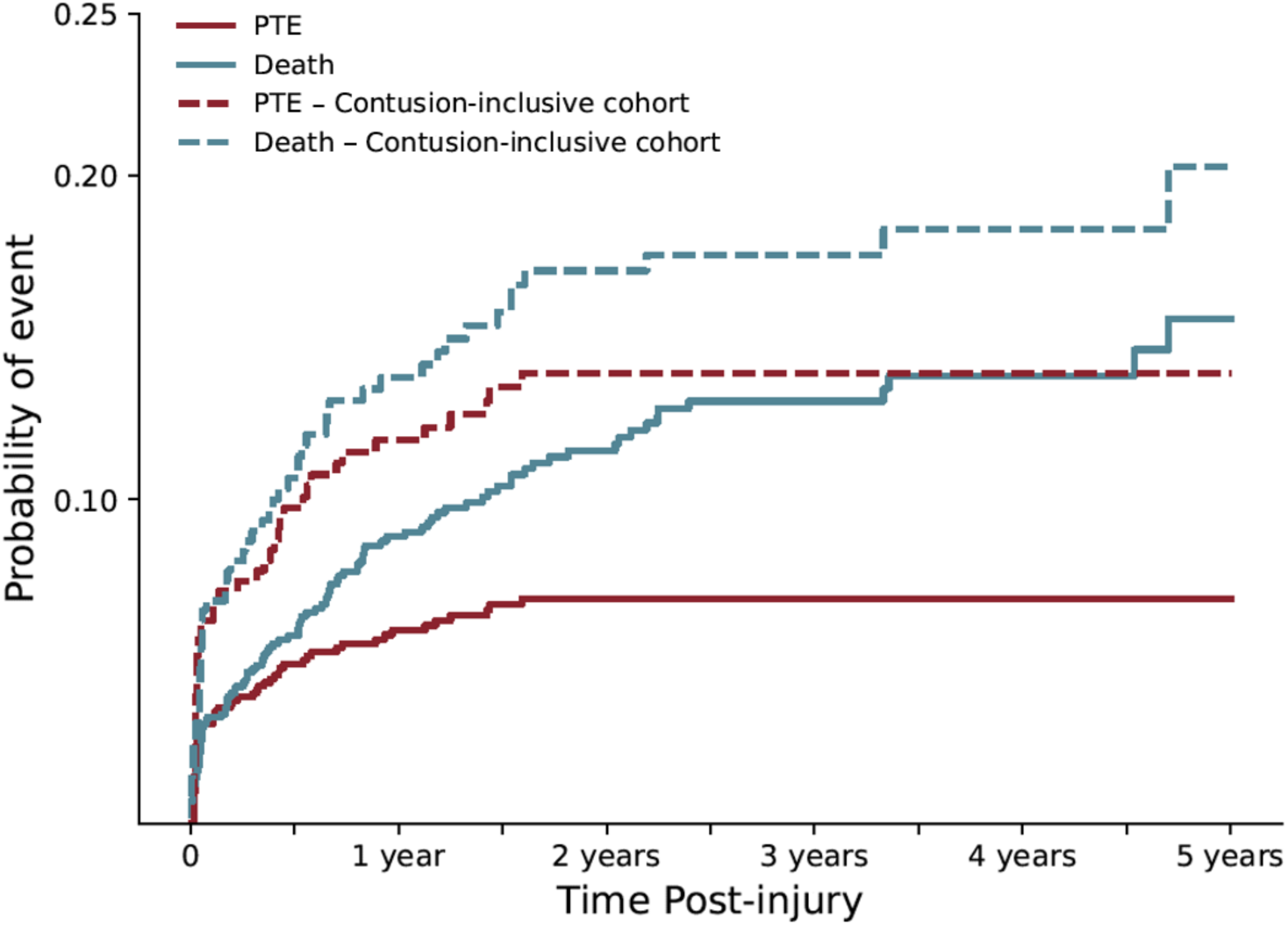
Cumulative incidences of post-traumatic epilepsy (PTE) and death over 5 years following traumatic brain injury (TBI). Solid lines show the full cohort used in the total hemorrhage and regional models, and dashed lines show the contusion-inclusive cohort used in sensitivity analyses. In the full cohort, the cumulative incidence of post-traumatic epilepsy (PTE) was 6% at 1 year, 6.9% at 2 years, and 6.9% at 5 years, while the cumulative incidence of death was 8.9%, 11.5%, and 15.6% at the same time points. In the contusion-inclusive cohort, the cumulative incidence of PTE was 11.8%, 13.9%, and 13.9% and the cumulative incidence of death was 13.8%, 17.1%, and 20.3% at 1, 2, and 5 years, respectively. At 5 years post-TBI, the difference in cumulative incidence between cohorts was 7% for PTE and 4.3% for death.

Consistent with the original total hemorrhage model, the sensitivity analysis for the PTE model retained Total Contusion Volume (HRΔ_5cc_ 1.14, 95% CI 1.07-1.19) as an independent predictor. For post-TBI mortality, Total Contusion Volume (HR_Δ5cc_ 1.07, 95% CI 1.002-1.15), IVH Volume (HR_Δ1cc_ 1.74, 95% CI 1.07-2.82), and EAH Volume (HR_Δ5cc_ 1.09, 95% CI 1.04-1.15) were significant independent predictors (Supplemental Table 13). Model performance was evaluated using the same methodology to the primary models. The PTE model achieved an average C-index of 0.794 and an overall mean AUC of 0.815, alongside a mean Brier score of 0.097. The mortality model performed with an average C-index of 746, an overall average AUC of 0.761, and a mean Brier score of 0.121 (Supplemental Table 9-12; Supplemental Figure 3C).

### Lesion Symptom Mapping Analysis

To map which contusion patterns predict outcomes the best, lesions symptom mapping through canonical correlation analysis was performed. For PTE, the resulting contusion map showed maximum weights localized at frontal insular and temporal regions (cross-validation r=0.27, P<0.001) (Figure 5). For mortality, while voxels with contusions collectively improved risk prediction (cross-validation r=0.11, P<0.001), no voxel clusters survived thresholding.

**Figure 5.**
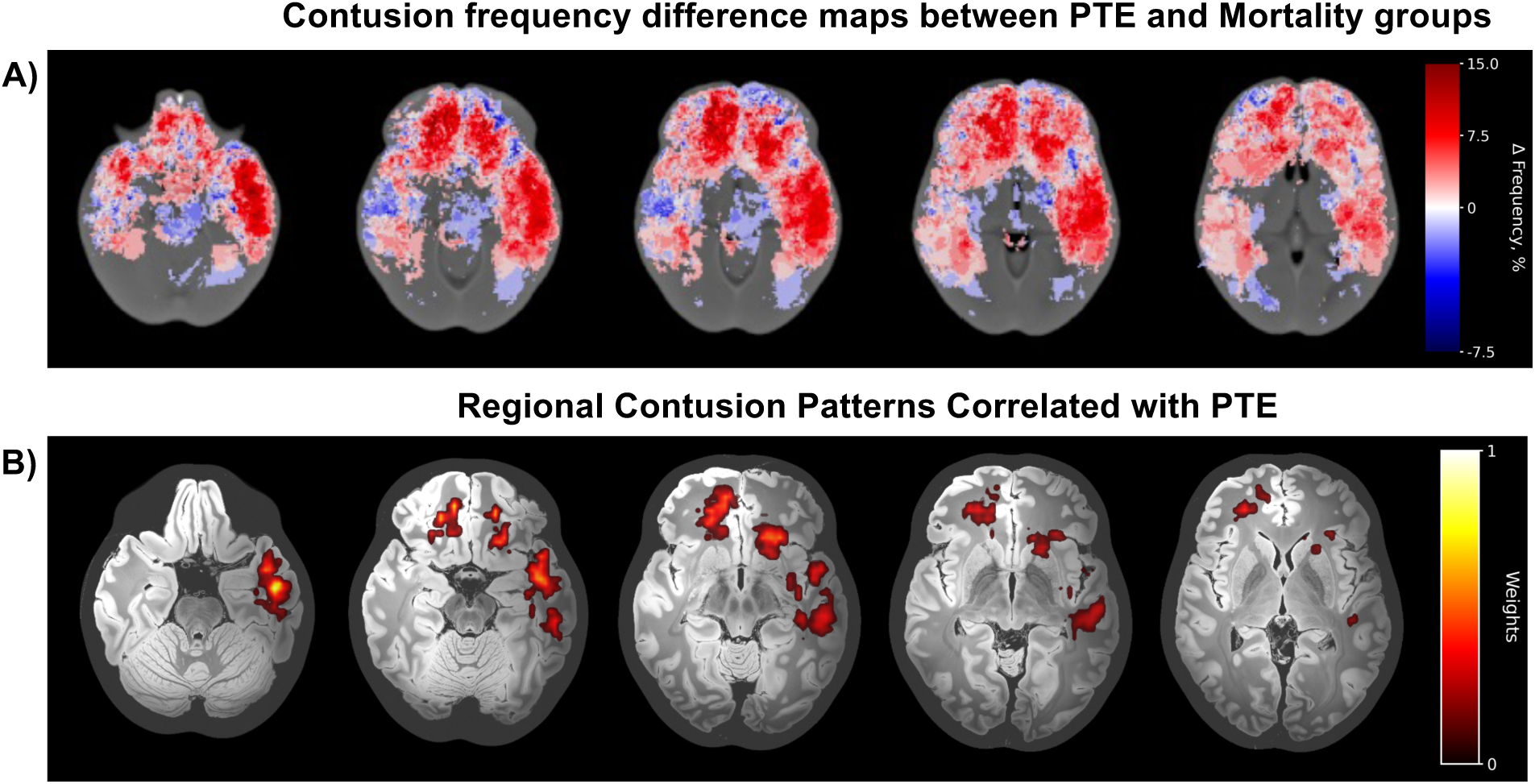
Spatial Analysis of Contusion Distribution and Outcome Associations A) Axial slices showing voxel-wise differences in contusion frequency (Δ Frequency, % = PTE − Mortality). Contusion masks were registered to a common template and averaged within each outcome group. Red indicates higher contusion frequency in PTE patients; blue indicates higher frequency in Mortality patients per voxel. Color scale: −7.5% to +15.0%, centered at zero. B) Predictive contusion patterns for PTE identified by sparse canonical correlation analysis. Color intensity represents normalized voxel weights (0-1) indicating association strength between contusion location and PTE outcome. Weights <10% of peak value set to zero. Analysis included voxels with lesions in ≥5 patients. (cross-validation r=0.27, P<0.001) z = -25, -18, -10, -5, 0 mm in MNI space. Please refer to Supplemental Figure 4 for further visualization.

## DISCUSSION

In this study, we investigated the utility of automated hemorrhagic volume measurements for prediction of PTE accounting for post-TBI mortality as a competing risk in a representative cohort of TBI patients. Hemorrhage volumes, along with demographic and clinical features, were associated with both outcomes in univariate analyses. In multivariable models using total hemorrhage measurements, contusion and EAH volumes were significantly associated with PTE and mortality. Furthermore, regionalized contusion volumes significantly predicted PTE, highlighting the role of lesion location in shaping long-term PTE risk after TBI. Our findings demonstrate that automated volumetric tools can be used to differentiate neuroimaging predictors of distinct outcomes and provide a framework for risk stratification of specific outcomes after TBI.

While previous studies had looked at PTE and post-TBI mortality risk factors separately, few if any have evaluated them together. Often PTE studies will eliminate patients who die from their study or simply censor them out.^13,27,43^ Using our competing risk framework our univariate analysis confirmed a wide range of predictors for PTE and mortality.^2,22,44,45^ Predictors of PTE included age, GCS score, post-traumatic seizures, and neurosurgical intervention within 7 days. We also found neuroimaging predictors such as contusion and EAH. Univariate post-TBI mortality predictors we found also corresponded to previous literature reports including age, admission GCS score, absent pupillary reaction, lower hemoglobin at admission, contusion, EAH and IVH.^24,46,47^

Many prior models did not account for mortality as a competing outcome, which may have confounded the observed relationship of contusion burden and PTE, conflating seizure risk and overall injury severity.^9^ In our multivariable analyses, contusion volume remained independently associated with both PTE and mortality. This reinforces that prior work linking contusions and EAH with PTE hold,^4,13,14,27^ even after accounting for mortality as a competing risk. By modeling mortality alongside PTE in a competing risk framework, with other injury severity factors, we can now provide more precise estimates of a contusion burden’s independent effect on PTE. To further assess the added value of automated imaging features beyond known clinical predictors, we compared the performance of our total hemorrhage model vs. a model using clinical features alone (Supplementary Tables 13-15). Incorporating imaging measures improved model discrimination and significantly enhanced calibration and accuracy of risk estimation (PTE: χ² = 29.4, p<0.001; Death: χ² = 21.6, p<0.001), indicating that quantitative imaging provides prognostic information that may improve real-world risk prediction in TBI beyond the clinical predictors.

Some previous PTE studies restricted their analyses to only severe TBI patients or limited their cohort to patients with contusions.^13,45^ Given TBI represents a broad spectrum of pathologies, ^23^ we used broader inclusion criteria that allowed assessment of patients with and without acute traumatic hemorrhages on CT.^13,14,27^ To compare how restricting analyses to only patients who have at least one contusion (“contusion-inclusive”) might yield different outcomes versus our broader cohort, we performed a sensitivity analysis. Notably, in the contusion-restricted cohort, the cumulative incidence of both PTE and mortality was higher than in the overall cohort, with a comparatively greater increase observed for PTE (Figure 4). These findings suggests that the contusion-inclusive subgroup may be a relatively more severe TBI subgroup than the general TBI population, irrespective of GCS.^12^ These findings also imply that lesion-targeted cohort selection may alter the observed estimations of PTE incidence and mortality competing risk assessments in the general TBI population.

Our regional investigations demonstrated contusion volumes are independently associated with PTE outcomes. Consistent with prior work, frontal and temporal regions are associated with increased risk of PTE.^16,17,26^ Previous work has also demonstrated that contusion location modulates outcome severity, with temporal contusions of comparable size more strongly associated with morbidity and mortality than frontal contusions.^34^ Our lesion heatmaps contrasted the varying frequencies of regional contusions between PTE and death outcomes (Figure 5).

Frontotemporal contusions were significantly associated with both outcomes, but more frequent among PTE patients; whereas brainstem contusions are more frequent in mortality, though did not reach statistical significance. Voxel-wise analysis identified contusion clusters at frontal, temporal, and insular as predictive of PTE. We did not find any clusters associated with mortality which is harder to capture with the inherently stricter thresholding of this analysis^40^ especially in small regions like the brainstem, or it may be related to the fact that TBI-related mortality is often diffuse or multicompartmental rather than focal. These regional findings exemplify how automated contusion tools could be leveraged to validate prior work, explore large-scale datasets for TBI endophenotypes and potentially be incorporated into a future workflows for prediction modeling of TBI outcomes.

TBI is inherently heterogeneous, and incorporating such heterogeneity in predictive modeling is essential but challenging. Automated hemorrhage measurements enabled us to conduct systematic investigations at larger scale than is typical for a single manual annotation project. However, several limitations must be considered. Additional use cases should be explored to optimize automated hemorrhagic measurements, as we identified a subset of patients with inaccurate segmentations in our analysis (Supplemental Fig. 1,2). Alternative algorithms or further training of current model may help improve segmentation accuracy. Furthermore, post-TBI mortality is influenced by factors beyond injury severity, especially decisions regarding withdrawal of life-sustaining therapy, meaning some mortality events may reflect goals of care decisions rather than underlying injury severity. Future studies might address this limitation through validation in cohorts with lower withdrawal-of-care rates or with sensitivity analyses restricted to mortality events less influenced by treatment limitations.

### Conclusions

Together, our findings suggest that, automatically measured contusion volumes from acute CT imaging could help predict post-traumatic epilepsy. By incorporating the competing risk of post-TBI mortality, we help define the associations of contusion volume and location on PTE and mortality outcomes in both patients with and without contusions. This work provides the foundation for future multi-center studies to further validate and explore its potential for clinical implementation.

## ACKNOWLEDGEMENTS

We would like to thank Yale University Library’s StatLab, specifically Atalay Demiray, MD, for consultation on statistical modeling. SBS receives support from the NIH/NINDS (K23NS136767). KNS receives support from the NIH/NINDS (UG3NS133209, U24NS129500, U01NS106513, U24NS107215, R21NS145048, R01EB031114) and the NIH/NIMHD (R01MD016178). BLE receives support from NIH/NINDS (R01NS138257). SFZ receives support from the National Institutes of Health (NIH R01NS126282, R01NS131347), and National Institute of Aging (NIA R01AG082693). AFS receives support from the National Institute of Neurological Disorders and Stroke (NINDS, R01NS111022, and R01NS126282). LJH receives support from the Daniel Raymond Wong Neurology Research Fund and the Yale NORSE/FIRES Biorepository Fund. EJG receives support from the NINDS (R01NS117904). MBW receives funding from the National Institutes of Health (NIH, RF1AG064312, RF1NS120947, R01AG073410, R01HL161253, R01NS126282, R01AG073598, R01NS131347, and R01NS130119), and the National Science Foundation (NSF, 2014431). JAK receives funding from the NINDS (K23NS112596-01A1, R01NS117904, R21NS128641, R01NS126282), the Brain Aneurysm Foundation and Swebilius Foundation.

## AUTHOR CONTRIBUTIONS

S.S., L.H., A.S., M.B.W., S.F.Z., A.F.S., K.N.S., S.B.O., B.L,E., E.J.G., and J.A.K. contributed to the conception and design of the article; B.B.A., J.R.W., D.S.J., J.A., V.T-L., N.S., W.S., D.D., K.T.C., J.K., and M.S. contributed to the acquisition and/or analysis of data; B.B.A. J.R.W. and J.A.K. contributed to drafting the text or preparing the figures.

## DATA AVAILABILITY

Analysis codes will be made publicly available upon publication at https://github.com/KimBAP-Lab. The data that support the findings of this study are not publicly available due to patient privacy and ethical restrictions. De-identified data are available to qualified researchers from the corresponding author upon reasonable request, and execution of an appropriate data use agreement.

