## Supplemental for "Predicting Post-Traumatic Epilepsy with Automated Contusion Measurements using Acute CT Images: A Competing Risk Approach"

Supplemental Figure 1: BLAST-CT Validation

Supplemental Table 1: IPH and Edema Measurements Univariate Correlations

Supplemental Figure 2: Study Cohort Identification Flowchart

Supplemental Table 2: Descriptive Analysis of Full Study Cohort

Supplemental Table 3: TBI Severity Subgroups Automated Measurement Comparison

Supplemental Table 4: Regional Contusion Measurement Comparison Across Outcome Groups

Supplemental Table 5,6: Model Performances/Calibration Scores for Total Hemorrhage Model

Supplemental Table 7,8: Model Performances/Calibration Scores for Regional Contusion Model

Supplemental Figure 3: Calibration Plots

Supplemental Figure 4: Contusion Topography Maps

Supplemental Table 9: Contusion-inclusive Model: Univariate Analysis

Supplemental Table 10,11,12c: Contusion-inclusive Model: Multivariate Analysis and Performance Metrics

Supplemental Table 13: Clinical Only Model: Multivariate Analysis

Supplemental Table 14,15: Clinical Only Model: Performance Metrics

**BLASTCT Validation**

We identified a retrospective cohort of patients admitted with Traumatic Brain Injury (TBI) between 2013-2021 across two tertiary care centers. Each PTE patient was matched with a single non-PTE patient based on: (1) TBI severity (a.k.a., admission Glasgow Coma Scale), age (±5 years), and sex. Identified 56 patients who developed PTE were case control matched with 56 non-PTE patients based on TBI severity (median GCS PTE=10, non-PTE=10, p=1.0), age (PTE=55, non-PTE=56, p=0.717), and sex (number female: PTE=16, non-PTE=26, p=0.053), for a total of 112 patients. We used Horos software (v3.3.1) for manual volume calculations. Two trained, blinded reviewers calculated volumes at each site (JRW, NS, JAK, WS). For each scan, reviewers traced the total circumference of hemorrhagic contusion and edema present on each slice. We compared manual and automated combined IPH and edema volumes using Pearson correlation.

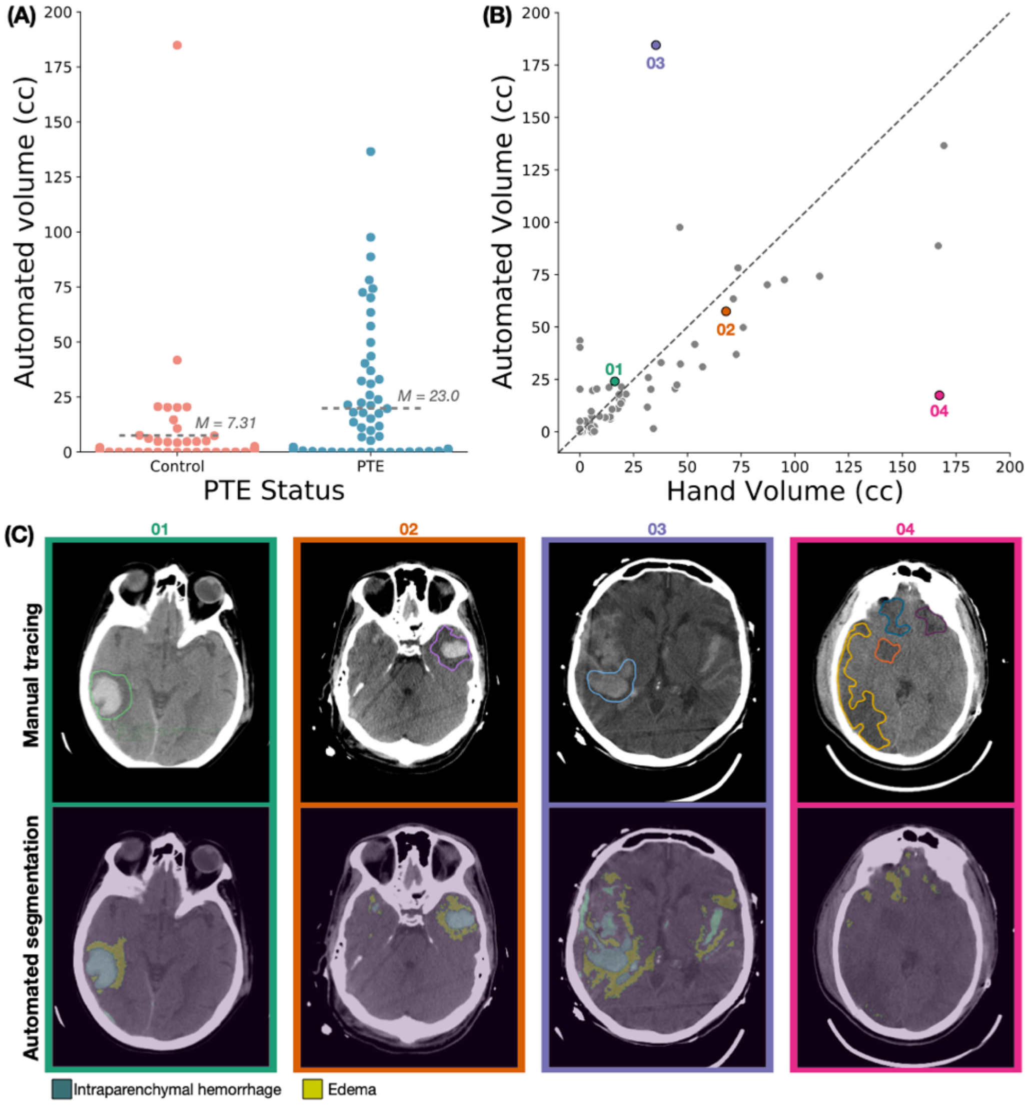

**Supplemental Figure 1.** Automated traumatic lesion volumes may be useful in prediction of PTE despite some limitations. (A) Automated volume is associated with PTE. Automated contusion volumes are shown for all control and PTE patients. PTE patients demonstrated increased contusion volume compared to controls. Each dot represents a single patient, and the dashed line represents the mean volume. (B) Automated and manual contusion volumes are correlated. A scatterplot demonstrating the automated volume compared to the manual volume is shown for each patient. Volumes were strongly correlated ($R=0.711$, $p<0.0001$). (C) Examples of manual and automated tracings are shown to demonstrate the ability of BLAST-CT to segment localized hemorrhage and perihematomal edema and highlight cases with large discrepancies. Scans correspond to the marked dots from the scatterplot in panel B.

**Intraparenchymal Hemorrhage (IPH) and Edema Measurements: Univariate Correlations**

Contusion volume (computed as IPH + edema) utilized as the primary metric for the multivariate analyses. IPH and edema measurements both total and regional are analyzed separately in univariate analysis for sensitivity analysis purposes.

**Supplemental Table 1.** Univariate cause-specific Cox proportional hazards models for PTE and post-TBI death: IPH and Edema Measurements

| | PTE | | Death | |
|---|---|---|---|---|
| Features | HR (95%CI) | p | HR (95%CI) | p |
| Total IPH | 1.07 (1.06-1.09) | **<0.001** | 1.04 (1.02-1.06) | **<0.001** |
| Total Edema | 1.06 (1.05-1.07) | **<0.001** | 1.04 (1.03-1.06) | **<0.001** |
| Frontal IPH | 1.09 (1.06-1.12) | **<0.001** | 1.06 (1.02-1.1) | **<0.001** |
| Temporal IPH | 1.14 (1.1-1.18) | **<0.001** | 1.05 (0.98-1.12) | 0.19 |
| Parietal IPH | 1.13 (1.03-1.24) | **0.009** | 1.12 (1.03-1.21) | **0.01** |
| Occipital IPH | 1.03 (0.77-1.39) | 0.827 | 1.06 (0.91-1.24) | 0.46 |
| Insular IPH | 2.15 (1.7-2.72) | **<0.001** | 1.75 (1.33-2.32) | **<0.001** |
| Basal Ganglia IPH | 1.18 (1.12-1.25) | **<0.001** | 1.06 (0.97-1.16) | 0.18 |
| Brainstem IPH | 3.92 (1.97-7.8) | **<0.001** | 1.83 (0.56-6.0) | 0.32 |
| Frontal Edema | 1.11 (1.07-1.15) | **<0.001** | 1.08 (1.04-1.11) | **<0.001** |
| Temporal Edema | 1.16 (1.12-1.2) | **<0.001** | 1.11 (1.06-1.17) | **<0.001** |
| Parietal Edema | 1.21 (1.1-1.33) | **<0.001** | 1.15 (1.04-1.28) | **0.01** |
| Occipital Edema | 1.13 (0.9-1.43) | 0.3 | 1.1 (0.92-1.32) | 0.27 |
| Insula Edema | 1.94 (1.55-2.42) | **<0.001** | 1.53 (1.21-1.94) | **<0.001** |
| Basal Ganglia Edema | 1.22 (1.14-1.32) | **<0.001** | 1.16 (1.09-1.25) | **<0.001** |
| Brainstem Edema | 1.28 (1.02-1.62) | **0.035** | 1.39 (1.18-1.64) | **<0.001** |

PTE = post-traumatic epilepsy; HR = hazard ratio; CI = confidence interval, IPH = intraparenchymal hemorrhage

**Study Cohort Identification**

**Supplemental Figure-2.** Study cohort selection flow diagram.
A total of 1,943 patients presenting to a tertiary trauma center between 2014 and 2025 met the initial inclusion criteria. Exclusions were applied for the following reasons: prior history of epilepsy or other acute brain injuries (n = 418), unusable or incomplete head CT scans (n = 285), and missing clinical features required for analysis (n = 363). After these exclusions, the final analysis cohort consisted of 1,017 patients who underwent automated lesion quantification and were included in predictive modeling. TBI = traumatic brain injury; AIS = acute ischemic stroke; ICH = intracerebral hemorrhage; CA = cardiac arrest; CT = Computed tomography.

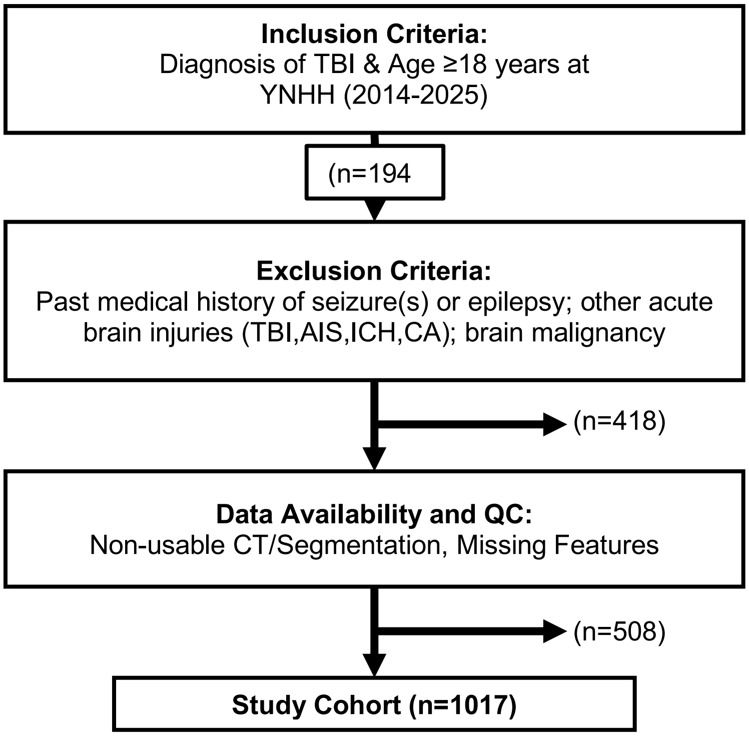

**Descriptive Analysis of the Study Cohort**

Continuous variables are presented as median [interquartile range (IQR)]. Categorical variables are presented as counts (n) and percentages (%). Normality was assessed using the Shapiro–Wilk test and homogeneity of variance with Levene's test. Depending on distributional assumptions, group comparisons were performed using one-way ANOVA with post hoc t-tests (parametric) or Kruskal–Wallis tests with pairwise Mann–Whitney U tests (nonparametric). Categorical variables were compared using $\chi^2$ tests. Pairwise p-values are reported only for features with overall statistical significance. Contusion volumes represent the sum of parenchymal intraparenchymal hemorrhage (IPH) and edema labels. Contusion prevalence was calculated by thresholding BLAST-CT derived measurements of IPH + edema at ≥1 cc.

**Supplemental Table-2** Baseline demographic, clinical, and imaging characteristics of the study cohort and outcome group comparisons.

| | | | | | | Pairwise Comparisons | | |
|---|---|---|---|---|---|---|---|---|
| Characteristics | Total (n=1017) | PTE (n=62) | No Event (n=848) | Death (n=107) | p-value | PTE vs No Event | PTE vs Death | Death vs No Event |
| Age at TBI, median (IQR), *year* | 63 (43.0-76.0) | 45 (28.0-66.5) | 62 (42.0-75.0) | 79 (66.0 - 86.0) | **<.001** | **<.001** | **<.001** | **<.001** |
| Sex, No. (%) | | | | | **0.01** | | | |
| Female | 390 (38.3%) | 14 (22.6%) | 329 (38.8%) | 47 (43.9%) | | **0.03** | **0.02** | **0.72** |
| Male | 627 (61.7%) | 48 (77.4%) | 519 (61.2%) | 60 (56.1%) | | **0.03** | **0.02** | **0.72** |
| Race, No. (%) | | | | | **0.002** | | | |
| American Indian or Native American | 1 (0.1%) | 0 (0.0%) | 1 (0.1%) | 0 (0.0%) | | N/A | N/A | N/A |
| Asian | 30 (2.9%) | 2 (3.2%) | 28 (3.3%) | 0 (0.0%) | | 1 | N/A | N/A |
| Black or African American | 119 (11.7%) | 13 (21.0%) | 96 (11.3%) | 10 (9.3%) | | 0.2 | 0.18 | 1 |
| Hispanic or Latino | 147 (14.5%) | 8 (12.9%) | 132 (15.6%) | 7 (6.5%) | | 1 | 0.52 | 0.06 |
| I Do Not Know | 8 (0.8%) | 0 (0.0%) | 8 (0.9%) | 0 (0.0%) | | N/A | N/A | N/A |
| I Do Not See My Race Listed Here | 8 (0.8%) | 2 (3.2%) | 5 (0.6%) | 1 (0.9%) | | 0.49 | 0.63 | 1 |
| I Prefer Not To Share | 2 (0.2%) | 0 (0.0%) | 2 (0.2%) | 0 (0.0%) | | N/A | N/A | N/A |
| Multi-Racial | 6 (0.6%) | 0 (0.0%) | 6 (0.7%) | 0 (0.0%) | | N/A | N/A | N/A |

| | | | | | | | | |
|---|---|---|---|---|---|---|---|---|
| Native Hawaiian or Other Pacific Islander | 1 (0.1%) | 0 (0.0%) | 1 (0.1%) | 0 (0.0%) | | N/A | N/A | N/A |
| Pacific Islander | 1 (0.1%) | 1 (1.6%) | 0 (0.0%) | 0 (0.0%) | | N/A | N/A | N/A |
| White | 694 (68.2%) | 36 (58.1%) | 569 (67.1%) | 89 (83.2%) | | 0.57 | **<.001** | **<.001** |
| Ethnicity, No. (%) | | | | | 0.18 | | | |
| Hispanic or Latino | 148 (14.6%) | 9 (14.5%) | 132 (15.6%) | 7 (6.5%) | | | | |
| I Do Not Know | 13 (1.3%) | 0 (0.0%) | 11 (1.3%) | 2 (1.9%) | | | | |
| I Prefer Not To Share | 9 (0.9%) | 1 (1.6%) | 8 (0.9%) | 0 (0.0%) | | | | |
| Not Hispanic or Latino | 847 (83.3%) | 52 (83.9%) | 697 (82.2%) | 98 (91.6%) | | | | |
| Admission GCS Score, No. (%) | | | | | **<.001** | | | |
| 13–15 (Mild TBI) | 823 (80.9%) | 32 (51.6%) | 712 (84.0%) | 79 (73.8%) | | **<.001** | **0.02** | **0.03** |
| 9–12 (Moderate TBI) | 59 (5.8%) | 6 (9.7%) | 46 (5.4%) | 7 (6.5%) | | 0.27 | 0.66 | 0.8 |
| 3–8 (Severe TBI) | 135 (13.3%) | 24 (38.7%) | 90 (10.6%) | 21 (19.6%) | | **<.001** | **0.02** | **0.03** |
| Injury Mechanism, No. (%) | | | | | **<.001** | | | |
| Accel/Decel. | 121 (11.9%) | 23 (37.1%) | 89 (10.5%) | 9 (8.4%) | | **<.001** | **<.001** | 0.72 |
| Direct Impact | 260 (25.6%) | 9 (14.5%) | 235 (27.7%) | 16 (15.0%) | | 0.1 | 1 | **0.02** |
| Fall > 3ft | 126 (12.4%) | 6 (9.7%) | 110 (13.0%) | 10 (9.3%) | | 0.58 | 1 | 0.72 |
| Fall From Ground Level | 505 (49.7%) | 24 (38.7%) | 409 (48.2%) | 72 (67.3%) | | 0.38 | **<.001** | **<.001** |
| Penetrating Injury | 5 (0.5%) | 0 (0.0%) | 5 (0.6%) | 0 (0.0%) | | | | |
| Blood Glucose at Admission, median (IQR), g/dL | 123 (106 - 152) | 144 (118.25 - 186) | 122.50 (105 - 149) | 126 (105.50 - 156) | **<.001** | **<.001** | **0.003** | 0.49 |
| Hemoglobin at Admission, median (IQR), g/dL | 13.40 (12.30 - 14.50) | 13.05 (12.03 - 14) | 13.60 (12.40 - 14.60) | 12.30 (10.85 - 13.25) | **<.001** | **0.03** | **<.001** | **0.001** |
| Pupillary Reaction at Admission, No. (%) | | | | | **0.03** | | | |
| Both | 929 (91.3%) | 54 (87.1%) | 783 (92.3%) | 92 (86.0%) | | 0.44 | 1 | 0.08 |
| None | 44 (4.3%) | 6 (9.7%) | 29 (3.4%) | 9 (8.4%) | | 0.1 | 1 | 0.08 |
| One | 44 (4.3%) | 2 (3.2%) | 36 (4.2%) | 6 (5.6%) | | 0.95 | 1 | 0.69 |

| | | | | | | | | |
|---|---|---|---|---|---|---|---|---|
| Posttraumatic Seizures <7 days, No. (%) | 75 (7.4%) | 21 (33.9%) | 47 (5.5%) | 7 (6.5%) | **<.001** | **<.001** | **<.001** | 1 |
| Neurosurgical intervention <7 days, No. (%) | 117 (11.5%) | 23 (37.1%) | 84 (9.9%) | 10 (9.3%) | **<.001** | **<.001** | **<.001** | 1 |
| Lesion Segmentation Prevalences, No. (%) | | | | | | | | |
| Contusion | 353 (34.7%) | 44 (71.0%) | 253 (29.8%) | 56 (52.3%) | **< 0.001** | **< 0.001** | **0.03** | **< 0.001** |
| IVH | 49 (4.8%) | 2 (3.2%) | 37 (4.4%) | 10 (9.3%) | 0.1 | | | |
| EAH | 461 (45.3%) | 48 (77.4%) | 342 (40.3%) | 71 (66.4%) | **< 0.001** | **< 0.001** | 0.2 | **< 0.001** |
| Total Lesion Volumes, median (IQR), cc | | | | | | | | |
| IPH | 0.05 (0 - 0.30) | 1.16 (0.05 - 10.09) | 0.03 (0 - 0.21) | 0.12 (0.03 - 1.57) | **<.001** | **<.001** | **0.003** | **<.001** |
| IVH | 0.03 (0 - 0.12) | 0.03 (0 - 0.11) | 0.03 (0 - 0.12) | 0.05 (0.01 - 0.22) | **0.008** | 0.82 | 0.057 | **0.006** |
| Edema | 0.79 (0.20 - 3.34) | 5.78 (1.20 - 16.26) | 0.61 (0.17 - 2.34) | 1.88 (0.39 - 9.18) | **<.001** | **<.001** | **0.02** | **<.001** |
| EAH | 0.37 (0.05 - 1.37) | 3.85 (0.58 - 12.03) | 0.29 (0.03 - 1.03) | 0.97 (0.19 - 4.84) | **<.001** | **<.001** | **0.015** | **<.001** |
| Contusion | 0.50 (0.09 - 1.95) | 6.12 (0.66 - 21.91) | 0.42 (0.08 - 1.38) | 1.07 (0.37 - 8.12) | **<.001** | **<.001** | **0.008** | **<.001** |

TBI = traumatic brain injury; PTE = post-traumatic epilepsy; IQR = interquartile range; GCS = Glasgow Coma Scale; g/dL = grams per deciliter; Accel./Decel. = acceleration/deceleration injury; CT = computed tomography; IPH = intraparenchymal hemorrhage; IVH = intraventricular hemorrhage; EAH = extra-axial hemorrhage; cc = cubic centimeters.

### TBI Severity Subgroups Automated Measurement Comparison

**Supplemental Table 3.** TBI Severity Subgroups: Automated Hemorrhagic Volume Measurements Comparison

| **Hemorrhage Type** | **Total** | **PTE** | **Death** | **No Event** | **p** | **PTE vs Death** | **PTE vs No Event** | **Death vs No Event** |
|---|---|---|---|---|---|---|---|---|
| Mild TBI | | | | | | | | |
| Contusion (*IPH +Edema*) | 0.42 [0.08–1.38] | 2.21 [0.40–13.79] | 0.77 [0.19–2.60] | 0.36 [0.06–1.19] | **<0.001** | 0.1 | **<0.001** | **<0.001** |
| IVH | 0.03 [0.00–0.12] | 0.03 [0.00–0.09] | 0.05 [0.01–0.12] | 0.03 [0.00–0.12] | 0.3161 | | | |
| EAH | 0.66 [0.18–2.61] | 5.78 [0.70–18.34] | 1.38 [0.26–4.32] | 0.59 [0.17–2.14] | **<0.001** | **0.02** | **<0.001** | **0.001** |
| IPH | 0.03 [0.00–0.21] | 0.48 [0.04–4.83] | 0.07 [0.01–0.52] | 0.02 [0.00–0.18] | **<0.001** | **0.03** | **<0.001** | **0.001** |
| Edema | 0.31 [0.03–1.09] | 1.37 [0.30–11.56] | 0.71 [0.12–1.75] | 0.25 [0.03–0.95] | **<0.001** | 0.09 | **<0.001** | **<0.001** |
| Moderate TBI | | | | | | | | |
| Contusion (*IPH +Edema*) | 0.72 [0.20–15.31] | 15.88 [7.17–21.60] | 15.73 [6.44–34.42] | 0.65 [0.13–3.85] | **0.007** | 0.94 | 0.1 | **0.002** |
| IVH | 0.03 [0.00–0.11] | 0.02 [0.01–0.03] | 0.29 [0.12–0.49] | 0.04 [0.00–0.10] | **0.02** | **0.02** | 0.34 | **0.013** |
| EAH | 1.27 [0.28–6.38] | 2.56 [1.29–4.41] | 10.47 [4.93–14.56] | 1.00 [0.23–5.21] | 0.07 | 0.07 | 0.64 | **0.025** |
| IPH | 0.10 [0.02–3.37] | 5.55 [1.28–10.09] | 3.22 [0.78–12.58] | 0.08 [0.01–0.89] | **0.01** | 0.95 | 0.06 | **0.012** |
| Edema | 0.64 [0.18–6.52] | 6.66 [4.51–10.45] | 11.56 [4.31–23.26] | 0.43 [0.07–3.14] | 0.004 | 0.36 | 0.09 | **0.002** |
| Severe TBI | | | | | | | | |
| Contusion (*IPH +Edema*) | 1.22 [0.38–7.63] | 12.31 [1.70–27.94] | 6.91 [0.81–23.20] | 0.70 [0.22–2.67] | **<0.001** | 0.38 | **<0.001** | **0.001** |
| IVH | 0.06 [0.01–0.26] | 0.04 [0.00–0.18] | 0.27 [0.02–0.48] | 0.05 [0.01–0.23] | 0.07 | | | |
| EAH | 1.71 [0.28–8.97] | 8.67 [1.89–16.19] | 6.28 [1.88–42.63] | 0.96 [0.15–5.35] | **<0.001** | 0.8 | **<0.001** | **<0.001** |
| IPH | 0.22 [0.02–1.69] | 2.53 [0.30–13.89] | 0.60 [0.19–4.11] | 0.08 [0.00–0.73] | **<0.001** | 0.29 | **<0.001** | **0.002** |
| Edema | 0.86 [0.21–4.31] | 5.50 [1.19–20.83] | 4.39 [0.40–11.95] | 0.55 [0.14–1.63] | **<0.001** | 0.52 | **<0.001** | **0.006** |

PTE = post-traumatic epilepsy, IPH = intraparenchymal hemorrhage; IVH = intraventricular hemorrhage; EAH = extra-axial hemorrhage.

**Supplemental Table 4.** Regional Contusion Measurement Comparison Across Outcome Groups

| Region | PTE | Death | No Event | p | PTE vs No Event | PTE vs Death | Death vs No Event |
|---|---|---|---|---|---|---|---|
| Frontal | 1.40 (0.19–7.14) | 0.35 (0.04–2.30) | 0.06 (0.00–0.46) | **<0.001** | **<0.001** | **0.02** | **<0.001** |
| Temporal | 0.71 (0.11–4.98) | 0.26 (0.02–1.10) | 0.05 (0.00–0.29) | **<0.001** | **<0.001** | **0.007** | **<0.001** |
| Parietal | 0.00 (0.00–0.23) | 0.00 (0.00–0.04) | 0.00 (0.00–0.00) | **<0.001** | **<0.001** | 0.95 | **<0.001** |
| Occipital | 0.00 (0.00–0.00) | 0.00 (0.00–0.00) | 0.00 (0.00–0.00) | **<0.001** | **<0.001** | 0.38 | **0.005** |
| Insula | 0.07 (0.00–1.02) | 0.00 (0.00–0.24) | 0.00 (0.00–0.00) | **<0.001** | **<0.001** | **0.009** | **<0.001** |
| Basal Ganglia | 0.11 (0.00–1.39) | 0.03 (0.00–0.54) | 0.00 (0.00–0.02) | **<0.001** | **<0.001** | 0.12 | **<0.001** |

**Multivariate Model Performance & Calibration**
Model performance metrics and calibration scores for prediction of PTE and post-TBI mortality.

**Supplemental Table 5**. Time Dependent Model Performance and Calibration Scores of Total Hemorrhage Model for prediction of PTE and post-TBI mortality.

| | PTE | | Death | |
|---|---|---|---|---|
| Time Point | AUC | Brier Score | AUC | Brier Score |
| 6 months | 0.809 | 0.044 | 0.78 | 0.051 |
| 1 year | 0.799 | 0.051 | 0.807 | 0.074 |
| 1.5 years | 0.787 | 0.059 | 0.808 | 0.084 |
| 2 years | 0.785 | 0.061 | 0.82 | 0.091 |
| 2.5 years | 0.778 | 0.061 | 0.81 | 0.101 |
| 3 years | 0.768 | 0.061 | 0.823 | 0.101 |
| Average | 0.787 | 0.056 | 0.808 | 0.084 |

AUC: Area under the receiver operating characteristic curve

**Supplemental Table 6**. Model performance across folds in cross validation of Total Hemorrhage Model for prediction of PTE and post-TBI mortality.

| Fold | PTE C-Index | Death C-Index |
|---|---|---|
| 1 | 0.849 | 0.814 |
| 2 | 0.826 | 0.758 |
| 3 | 0.813 | 0.791 |
| 4 | 0.752 | 0.846 |
| 5 | 0.756 | 0.715 |
| Average | 0.799 | 0.785 |

C-index: concordance index

**Supplemental Table 7**. Time Dependent Model Performance and Calibration Scores of Regional Contusion Model (Model 2) for prediction of PTE and post-TBI mortality.

| | PTE | | Death | |
|---|---|---|---|---|
| Time Point | AUC | Brier Score | AUC | Brier Score |
| 6 months | 0.818 | 0.045 | 0.772 | 0.052 |
| 1 year | 0.809 | 0.052 | 0.801 | 0.075 |
| 1.5 years | 0.799 | 0.059 | 0.803 | 0.085 |
| 2 years | 0.795 | 0.061 | 0.817 | 0.092 |
| 2.5 years | 0.787 | 0.061 | 0.810 | 0.102 |
| 3 years | 0.774 | 0.062 | 0.822 | 0.102 |
| Average | 0.797 | 0.057 | 0.804 | 0.084 |

AUC: Area under the receiver operating characteristic curve

**Supplemental Table 8**. Model performance across folds in cross validation of Regional Contusion Model (Model 2) for prediction of PTE and post-TBI mortality.

| Fold | PTE C-Index | Death C-Index |
|---|---|---|
| 1 | 0.861 | 0.812 |
| 2 | 0.837 | 0.771 |
| 3 | 0.821 | 0.784 |
| 4 | 0.764 | 0.836 |
| 5 | 0.761 | 0.696 |
| Average | 0.809 | 0.78 |

C-index: concordance index

### Calibration Plots

Supplemental Figure 3. (A) Total Hemorrhage Model (Model 1) applied to the full TBI cohort. (B) Regional Contusion Model (Model 2) incorporating regionalized contusion volumes with total EAH and IVH volumes. (C) Sensitivity analysis restricted to patients with detectable contusions on BLAST-CT. Calibration plots comparing predicted absolute risk with observed event incidence across quantiles of predicted risk across all time points.

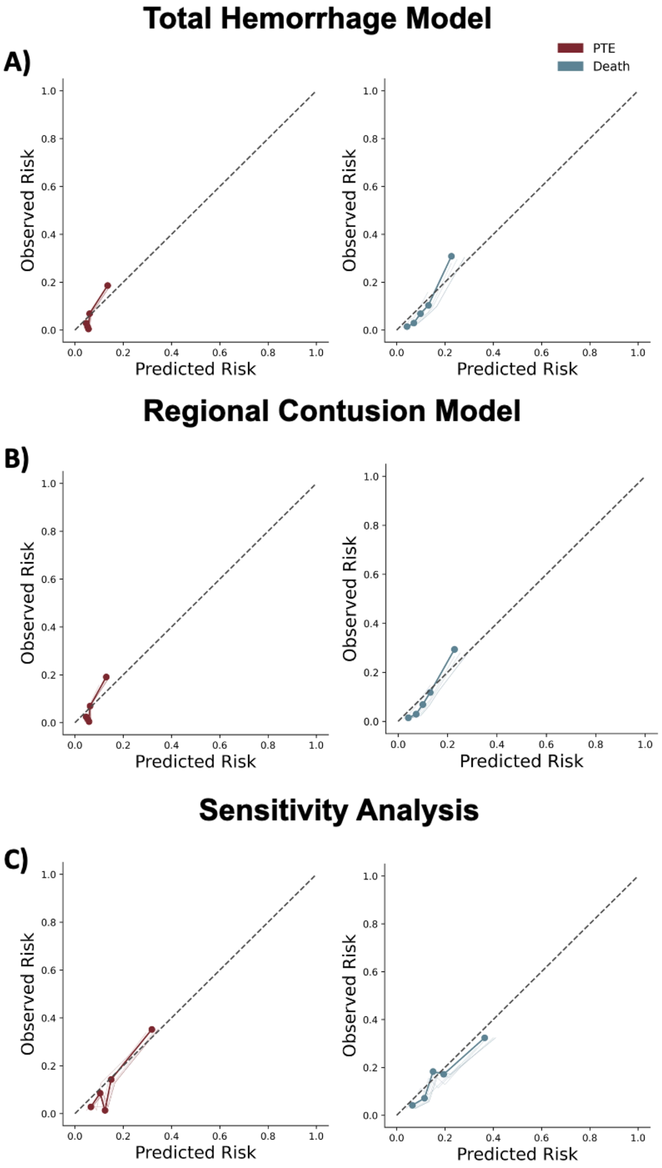


### Contusion Topography Maps

**Supplemental Figure 4-A**. Group-level difference map comparing the spatial distribution of contusions between patients who developed post-traumatic epilepsy (PTE) and those who died (Δ frequency, %).

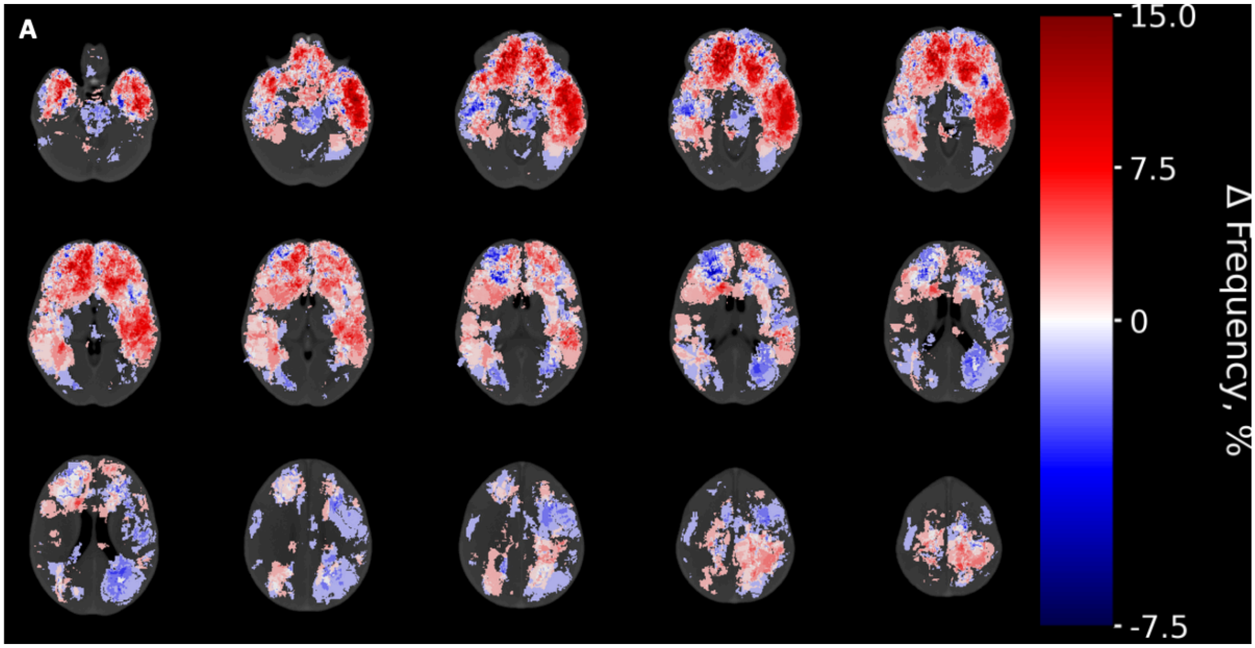


**Supplemental Figure 5-B**. Sparse canonical correlation analysis (SCCAN) lesion–outcome map showing voxelwise weights linking lesion location to PTE outcome.

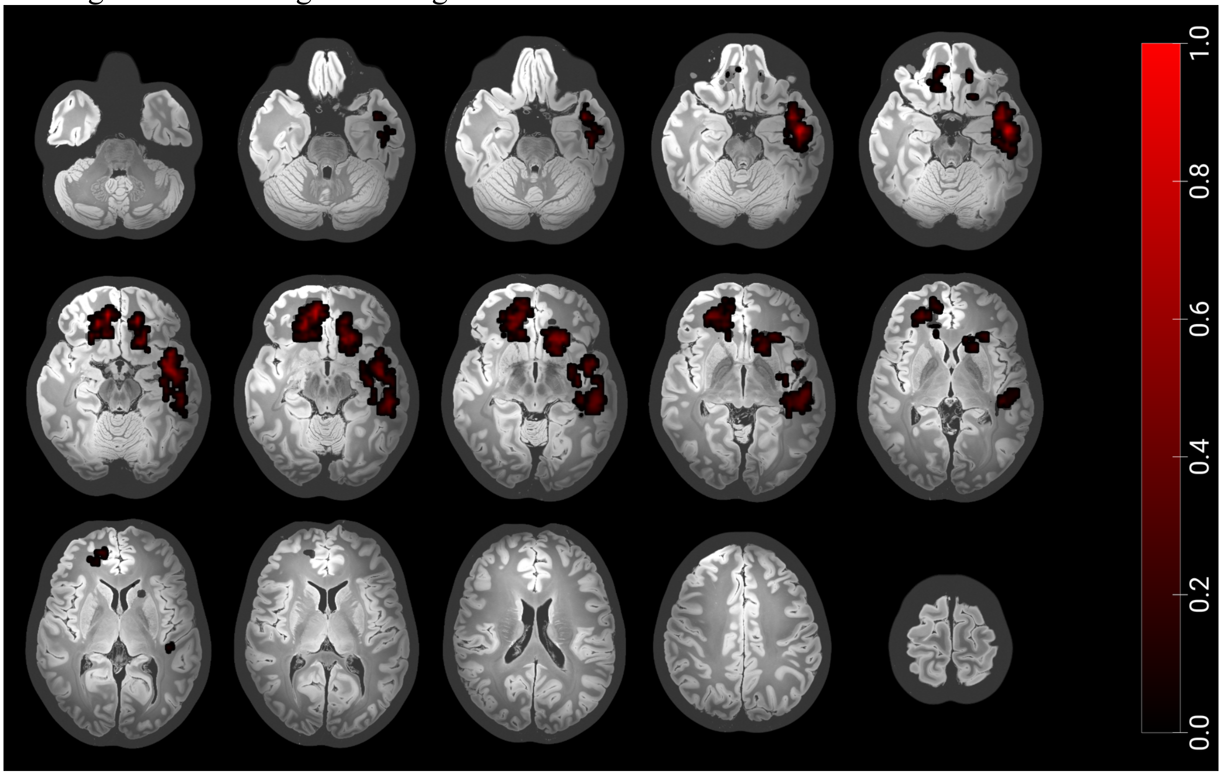


**Contusion-inclusive Cohort: Sensitivity Analysis Development**

Sensitivity analysis was performed after restricting the cohort to patients with contusion present, either in isolation or in combination with other hemorrhagic lesions. Patients with contusions were identified by applying a ≥1 cc threshold to IPH and edema masks across the full cohort. A cohort of 353 patients with contusions was identified, either in isolation or accompanied by other hemorrhagic lesions. Multivariable cause-specific Cox models were then developed in this subgroup to assess the impact of focusing analyses on contusion-inclusive populations, as has been done in prior PTE prediction literature.

**Supplemental Table 9.** Sensitivity Analysis: Univariate cause-specific Cox proportional hazards models for PTE and post-TBI mortality

| | PTE | | Death | |
|---|---|---|---|---|
| Features | HR (95%CI) | p | HR (95%CI) | p |
| Demographics | | | | |
| Age | 0.97 (0.96–0.99) | **<0.001** | 1.04 (1.02–1.06) | **<0.001** |
| Sex (Male) | 1.47 (0.71–3.06) | 0.303 | 0.80 (0.46–1.40) | 0.437 |
| Clinical Features | | | | |
| Total GCS | 0.86 (0.81–0.91) | **<0.001** | 0.94 (0.9-0.98) | **0.02** |
| GCS Eye Score | 0.57 (0.46–0.71) | **<0.001** | 0.83 (0.71–0.97) | **0.02** |
| GCS Verbal Score | 0.65 (0.55–0.77) | **<0.001** | 0.84 (0.68–1.03) | 0.096 |
| GCS Motor Score | 0.72 (0.63–0.82) | **<0.001** | 0.86 (0.75–0.99) | **0.034** |
| Pupillary Reaction (ref. Both) | | | | |
| One | 0.43 (0.06–3.10) | 0.400 | 0.68 (0.16–2.78) | 0.588 |
| None | 2.18 (0.78–6.09) | 0.139 | 3.43 (1.55–7.60) | **0.002** |
| Injury Mechanism (ref. GLF) | | | | |
| Accel/Decel | 5.30 (2.92–9.64) | **<0.001** | 0.85 (0.37–1.99) | 0.715 |
| Direct impact | 0.30 (0.11–0.84) | **0.022** | 0.41 (0.19–0.91) | **0.029** |
| Fall > 3ft | 0.76 (0.27–2.14) | 0.608 | 1.33 (0.63–2.81) | 0.457 |
| Penetrating injury | NA | 0.995 | NA | 0.99 |
| Blood Glucose at Admission | 1.00 (1.00–1.01) | **0.004** | 1.0 (1.0-1.0) | 0.77 |
| Hemoglobin at Admission | 0.96 (0.82–1.13) | 0.628 | 0.76 (0.68-0.86) | **<0.001** |
| Posttraumatic Seizures <7 days | 6.51 (3.55–11.93) | **<0.001** | 1.17 (0.50–2.74) | 0.713 |
| Neurosurgical Intervention <7 days | 3.46 (1.91–6.28) | **<0.001** | 0.76 (0.36–1.62) | 0.483 |
| Neuroimaging Variables | | | | |
| Total Segmentation Volumes | | | | |
| $IPH_{\Delta 5cc}$ | 1.30 (1.19–1.42) | **<0.001** | 1.12 (0.99–1.26) | 0.077 |
| $IVH_{\Delta 1cc}$ | 0.73 (0.24–2.23) | 0.581 | 1.80 (1.22–2.66) | **0.003** |
| $EAH_{\Delta 5cc}$ | 1.09 (1.03–1.16) | **0.003** | 1.13 (1.09–1.17) | **<0.001** |
| $Edema_{\Delta 5cc}$ | 1.24 (1.14–1.35) | **<0.001** | 1.18 (1.08–1.29) | **<0.001** |

| | | | | |
|---|---|---|---|---|
| Contusion (IPH+Edema) $_{\Delta 5cc}$ | 1.15 (1.10–1.21) | **<0.001** | 1.09 (1.03–1.15) | **0.003** |

Hazard ratios (HR) with 95% confidence intervals (CI) and p-values are reported. Hazard ratios (HR) with 95% confidence intervals (CI) and p-values are reported. PTE = post-traumatic epilepsy; HR = hazard ratio; CI = confidence interval; GCS = Glasgow Coma Scale; GLF = ground-level fall; IPH = intraparenchymal hemorrhage; IVH = intraventricular hemorrhage.

**Supplemental Table 10.** Multivariable cause-specific Cox regression models for PTE and post-TBI mortality in the sensitivity analysis cohort.

| Features | Coefficient (95%CI) | p-value |
|---|---|---|
| | PTE | |
| Posttraumatic Seizures <7 days | 3.62 (1.89-6.95) | **<0.001** |
| Total Contusion Volume $_{\Delta 5cc}$ | 1.14 (1.07-1.19) | **<0.001** |
| Total GCS | 0.88 (0.82-0.93) | **<0.001** |
| | Death | |
| IVH Volume $_{\Delta 1cc}$ | 1.74 (1.07-2.82) | **0.02** |
| Pupillary Reaction: None | 3.51 (1.26-9.76) | **0.01** |
| EAH Volume $_{\Delta 5cc}$ | 1.09 (1.04-1.15) | **<0.001** |
| Total Contusion Volume $_{\Delta 5cc}$ | 1.07 (1.002-1.15) | **0.04** |
| Age | 1.05 (1.03-1.07) | **<0.001** |
| Total GCS | 0.90 (0.83-0.97) | **0.009** |
| Hemoglobin | 0.79 (0.68-0.91) | **0.001** |

PTE = post-traumatic epilepsy; HR = hazard ratio; CI = confidence interval; GCS = Glasgow Coma Scale; GLF = ground-level fall; EAH = extra-axial hemorrhage; IVH = intraventricular hemorrhage.

**Supplemental Table 11.** Time Dependent Model Performance and Calibration Scores of Contusion-inclusive Cohort

| | PTE | | Death | |
|---|---|---|---|---|
| Time Point | AUC | Brier Score | AUC | Brier Score |
| 6 months | 0.826 | 0.077 | 0.727 | 0.091 |
| 1 year | 0.834 | 0.089 | 0.744 | 0.112 |
| 1.5 years | 0.820 | 0.102 | 0.765 | 0.125 |
| 2 years | 0.813 | 0.104 | 0.762 | 0.135 |
| 2.5 years | 0.807 | 0.104 | 0.778 | 0.133 |
| 3 years | 0.790 | 0.106 | 0.792 | 0.133 |
| Average | 0.815 | 0.097 | 0.761 | 0.121 |

AUC: Area under the receiver operating characteristic curve

**Supplemental Table 12.** Model performance across folds in cross validation of contusion-inclusive cohort

| Fold | PTE C-Index | Death C-Index |
|---|---|---|
| 1 | 0.789 | 0.642 |
| 2 | 0.761 | 0.806 |
| 3 | 0.879 | 0.748 |
| 4 | 0.809 | 0.805 |
| 5 | 0.731 | 0.73 |
| Average | 0.794 | 0.746 |

C-index: concordance index

**Assessment of Added Value of Imaging Features**

Clinical-only Model Development and Comparison: A clinical-only Cox model was built using the same cross-validated feature-selection procedure as the primary analysis, restricting the predictor pool to age, total GCS, glucose, hemoglobin, pupillary reactivity, mechanism-of-injury indicators, early seizures, surgery status, and sex. Model discrimination and calibration were evaluated using cross-validated concordance index, time-dependent AUC, and Brier scores. Nested likelihood ratio tests were then used to compare the clinical-only models with the corresponding total-hemorrhage models. For PTE, the full model showed a higher log-likelihood than the clinical-only model (LR $\chi^2 = 29.50$, df = 1, $p < 0.001$). For mortality, the full model likewise showed improved fit (LR $\chi^2 = 21.63$, df = 2, $p < 0.001$). Full model coefficients and cross-validation performance metrics for the clinical-only analysis are provided in the accompanying tables.

**Supplemental Table 13.** Multivariable cause-specific Cox regression models for PTE and post-TBI mortality using only the Clinical features.

| Features | Coefficient (95%CI) | p-value |
|---|---|---|
| | PTE | |
| Posttraumatic Seizures <7 days | 4.64 (2.66-8.09) | **<0.001** |
| Neurosurgical Intervention <7 days | 2.38 (1.32-4.28) | **0.003** |
| Total GCS | 0.88 (0.82-0.93) | **0.001** |
| | Death | |
| Age | 1.06 (1.04-1.07) | **<0.001** |
| Total GCS | 0.76 (0.69-0.84) | **<0.001** |
| Hemoglobin | 0.76 (0.69-0.84) | **<0.001** |

**Supplemental Table 14.** Time Dependent Model Performance and Calibration Scores of Clinical-only Models

| | PTE | | Death | |
|---|---|---|---|---|
| Time Point | AUC | Brier Score | AUC | Brier Score |
| 6 months | 0.801 | 0.046 | 0.742 | 0.053 |
| 1 year | 0.782 | 0.055 | 0.788 | 0.076 |
| 1.5 years | 0.761 | 0.063 | 0.791 | 0.087 |
| 2 years | 0.760 | 0.065 | 0.809 | 0.094 |
| 2.5 years | 0.756 | 0.065 | 0.807 | 0.103 |
| 3 years | 0.745 | 0.065 | 0.822 | 0.102 |
| Average | 0.768 | 0.060 | 0.793 | 0.086 |

**Supplemental Table 15.** Model performance across folds in cross validation of Clinical-only Models.

| Fold | PTE C-Index | Death C-Index |
|---|---|---|
| 1 | 0.745 | 0.815 |
| 2 | 0.862 | 0.749 |
| 3 | 0.755 | 0.74 |
| 4 | 0.728 | 0.826 |
| 5 | 0.782 | 0.67 |
| Average | 0.775 | 0.76 |
